# Evaluation of a Biplanar Vector-Based Diagnostic Model for Subthreshold Disease: In Silico Stress Testing of Composite Drift Score Performance Under Stochastic, 2D, and 3D Conditions

**DOI:** 10.1101/2025.06.19.25329940

**Authors:** Gaurav Prakash

## Abstract

Traditional threshold-based diagnostics often miss early disease progression, particularly in subthreshold states. In a prior model, we proposed the Composite Drift Score (CDS), a vector-based index that quantifies directional drift from physiological norms. This metric uses two components: the Magnitude-to-Noise Ratio (MNR) and a Directional Emphasis Multiplier (DEM). This study stress-tests the CDS model under a range of biologically plausible and extreme conditions using in silico synthetic data. A total of 2000 simulated subjects were generated with varying prevalence, pause patterns, intra-subject variability, and progression speed. Two versions of CDS were evaluated: one using Coefficient of Repeatability (CR) and another using Mahalanobis distance (MD). Model performance was quantified using the area under the curve for lead time (AUC-LT), reflecting both timing and detection advantage. Results showed that CDS outperformed threshold-based methods in early detection, particularly under fragmented or physiological-range noisy conditions. The Mahalanobis-based variant demonstrated higher resilience under stress. The framework was also expanded to a three-variable version, preserving directional behavior and performance characteristics, suggesting dimensional scalability. These in silico findings indicate that the CDS model may warrant further investigation in clinical datasets.

## Introduction

In a recent study, we proposed a biplanar model to distinguish disease and normal states to diagnose subthreshold disease. [1] This geometric framework was constructed on the premise that disease modeling in the subthreshold space can be performed using the magnitude of significant change and the alignment of the subject’s vector with a disease state vector. It uses the well-established concept of the tipping point to define drift, a non-self-correcting change from homeostasis [2,3]. We called this phenomenon ‘intentional directionality’. Compared to threshold-based diagnostic systems, this method does not wait for the breach of an arbitrary decision boundary and detects angular deviation from the physiological plane. The unitless metric used to quantify this change was termed Composite Drift Score (CDS), which is a customizable metric derived from 2 entities, the subject vector’s Magnitude to ‘physiological’ Noise Ratio (MNR) and Directional Emphasis Multiplier (DEM, a derivative of cosine between the subject vector and disease vector). In that initial exploratory study based on synthetic data based on population-based norms and induced stochasticity, the three indices (CDS, MNR, and DEM) were able to mimic the behavior of early progressive disease in the sub-threshold progressive cases, a cohort that is missed by the classic threshold model. In addition, these indices were stable in the stable normal and stable disease groups. Encouraged by these results, the present follow-up study aims to stress test the CDS framework under increasingly stochastic conditions and to extend the model to trivariate (3D) input data. This study is the next step in evaluating the robustness, dimensional scalability, and diagnostic behavior of the CDS index in silico. Stress testing the model under extreme and variable scenarios helps define its operating limits, clinical robustness, and interpretability across conditions.

## Methods

### 1. Existing framework

For this follow-up study, we build upon the geometric framework previously described in the index study to derive the CDS. We have supplied the framework as **Appendix A** in this study for easier review [1].

### 2. Synthetic dataset design

To evaluate the robustness, generalizability, and performance of the CDS index in comparison to the threshold-based tests, we designed a synthetic stress testing module. The module consisted of 2,000 new unlabeled computer-generated synthetic subjects, each with baseline values within the physiological (normal) range. Follow-up was simulated for 6 monthly follow-ups over a 10-year period to simulate a long-term follow-up.

Unlike the datasets created for the exploratory study, this was highly customizable. The base layer consisted of the two options generating a random patient which could be either stable (normal range baseline value + variable noise) or progressive (normal range baseline value + variable noise + variable drift) which was to be decided by computer-generated random numbers. The second layer was the customization to increase stochasticity further which allowed the following simulation options parameters, which could be varied independently and in combination across multiple scenarios by the user. All scenarios were linearly alterable. All simulations were implemented using Microsoft Excel for transparency and reproducibility at this early stage **(The CDS simulation framework, including the interactive dashboard, bivariate and trivariate models, and Excel modules, is available upon request for academic, non-commercial use only. These tools are not released under open access, and reuse or reproduction without explicit permission is not permitted).** All the options and functionality are visualized in the screenshot of the dashboard in **Figure 1**.

A. Prevalence Percentage (0–100%): this option alters the percentage of the subjects out of 2000 that are pre-decided to worsen. As the prevalence of a disease can affect threshold-based tests, we wanted to evaluate the effect in this comparison.
B. Visit-Level Stochasticity (Pause Probability, 0–100%): Even if a subject is on the path of pre-threshold worsening, there can be periods where a plateau is achieved for some time. These stable phases are very commonly seen in clinical situations and affect the rate of change-based tests. Our exploratory study’s cohort was only meant to account for a variable rate of change at each visit. This additional layer introduces a randomly selected absolute pause for that visit.
C. Increased **normal** eyes intrameasurement standard deviation (Sw inflation, no limits, normal group noise): The coefficient of repeatability (2.77x Sw) was used as the measure of noise when constructing the MNR metric. In real-life situations, we expect the noise in the pre-threshold cases to be like that of normal cases as currently, they come from the same pooled labeled as normal in population-based studies. However, to induce more customization (if the normal and disease populations are tested in different environments) we retained the capability to change the Sw for the normal group separately. This aimed to increase the noise value (denominator in the MNR metric) independently.
D. Increased test eyes intrameasurement standard deviation (disease group Sw inflation, no limits, test group noise): This aimed to increase the intrameasurement standard deviation in the test eyes independent of the normal eye.
E. Variable level drift rate change, no limits: Controls the rate of progression independently for all the variables (*x,y* in the 2D model and *x,y,z* in the 3D model). This allows testing of CDS performance if one or more variables are faster or slower than the population norm. One criticism of threshold-based models is that as they are created at a large population level, the pooled data does not account for subgroup customizations. For example, a sub-group may have a variable worsening faster than the global rate, and therefore, they may have false positive triggers, or vice versa. This customization serves to address that detail.
F. Custom Threshold Definition (X and Y axes): This allows users to manually set diagnostic thresholds for each variable. This simulates guideline updates or center-specific boundary shifts and helps test how CDS adapts to different diagnostic cutoffs.
G. Suspect or Borderline Zone (Custom Percentile Band): the definition of a suspect or borderline case has been the Achilles heel for threshold-based definitions. They define suspect as a function of the proximity to the division boundary, in the “gray zone” between normal and disease thresholds. This customization in the module allows the user to adjust this zone as a percentile of the difference between the mean normal value and the disease threshold cutoff. This enables finer diagnostic granularity and simulates changes in suspect zone criteria across studies or classification systems.
H. Three-dimensional expansion: in this stand-alone module, we expand the entire system to a three-variable system. The above customizations (1-6) are available both at the cohort and the variable level. In addition, we compare the MNR (and thus the derived metric CDS) derived from using CR and Mahalanobis distance with the conventional thresholds. We briefly explored the concept of using Mahalanobis distance in the exploratory study. [4,5]

**Figure 1.**
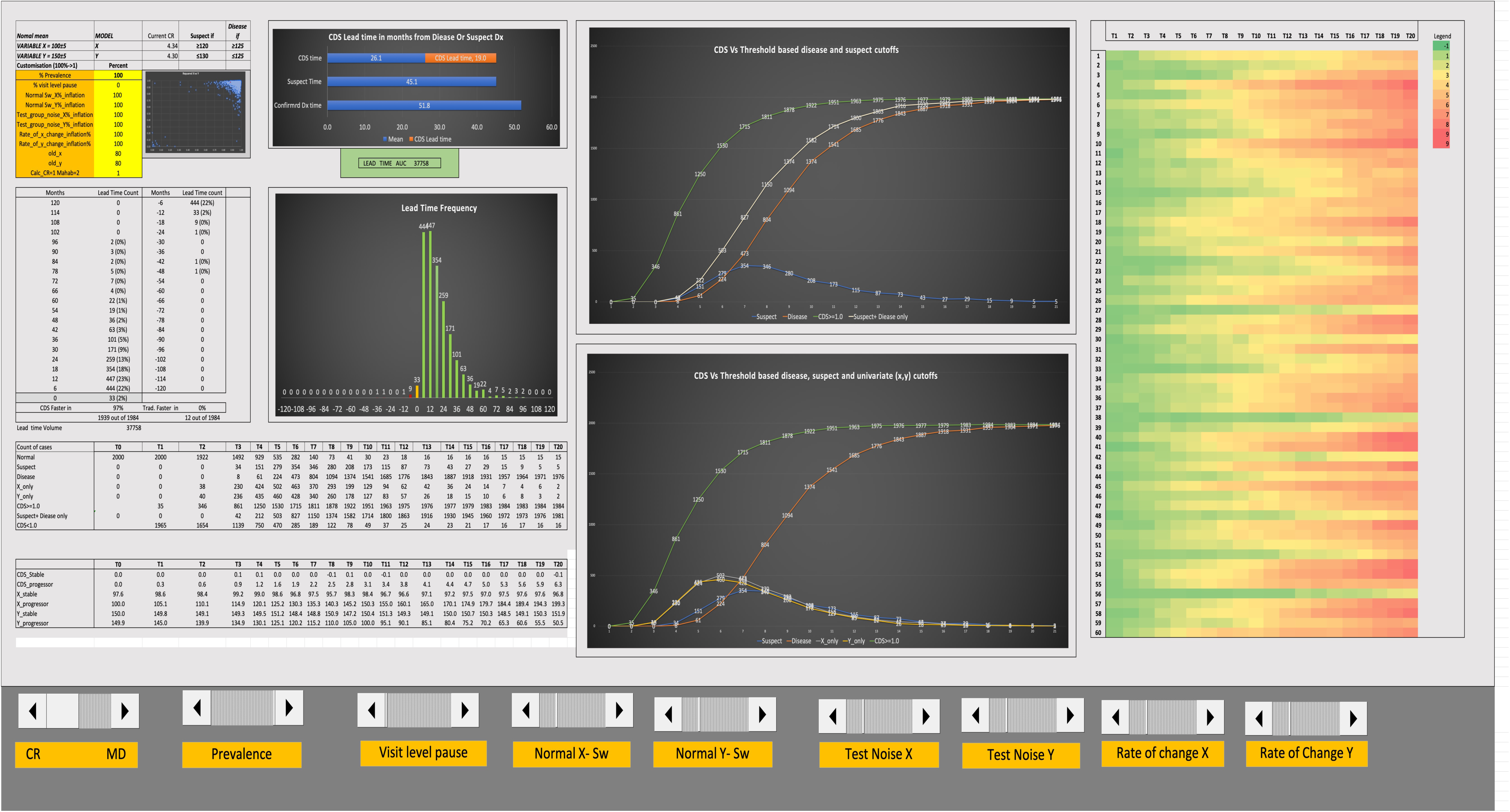
Standardized input/output panel for stress test scenarios in this study. Each panel presents a complete visualization of CDS system behavior for one simulation, with all key diagnostics arranged consistently. **Top-left**: summary of the scenario parameters, including disease prevalence, visit-level pause, Sw inflation, and rate of change. Fields highlighted in **yellow** are user-editable; others are locked or derived values. T**op-center**: horizontal bar chart displays the mean lead time (orange) for CDS, compared to suspect and threshold-based triggers. The green box below it shows the value of the Area under curve for lead time (positive values-AUC-LT higher for CDS, negative values-AUC-LT higher for cutoff based tests). **Far**-**right**: heatmap showing CDS signal for the first 60 test subject, with time on the x-axis and patient ID on the y-axis, with warmer colors showing higher CDS scores (color scale auto-adjusts to that particular simulation’s range of data). **Right of center-upper line graph** shows cumulative detection counts over time for CDS, suspect, and disease cutoffs, allowing visual comparison of signal lead time. **Right of center-lower line graph:** a second graph additionally plots individual variable cutoffs (X and Y) alongside CDS, suspect and disease thresholds, enabling interpretation of whether signal is univariate or multivariate. **Lower-right table**: trigger timepoints and classification status for each patient and serves as the data source for the adjacent plots. **Lower center**: histogram shows the frequency distribution of CDS lead times across the cohort, showing negative lead times in red and positive lead times in blue. **Lower-left section** summarizes average CDS, X, and Y values across stable and progressive subgroups. **Bottom toolbar** contains interactive controls for switching between CDS models (CR or Mahalanobis) and adjusting simulation parameters such as prevalence, noise, and pause rate.

**Figure 2:**
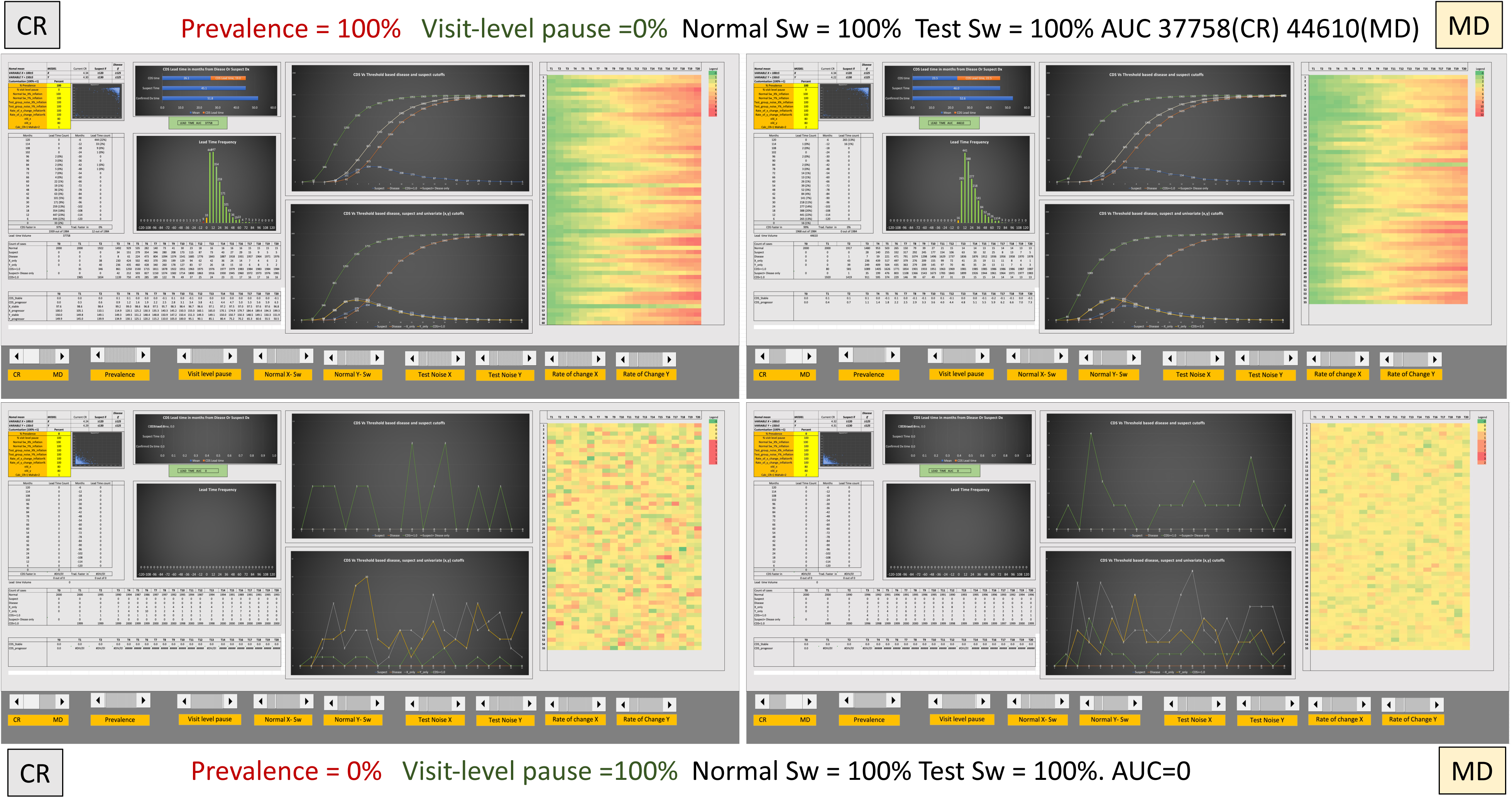
Screenshot of the performance of the CR and MD based frameworks on base case scenarios: full active and fully paused.

**Figure 3.**
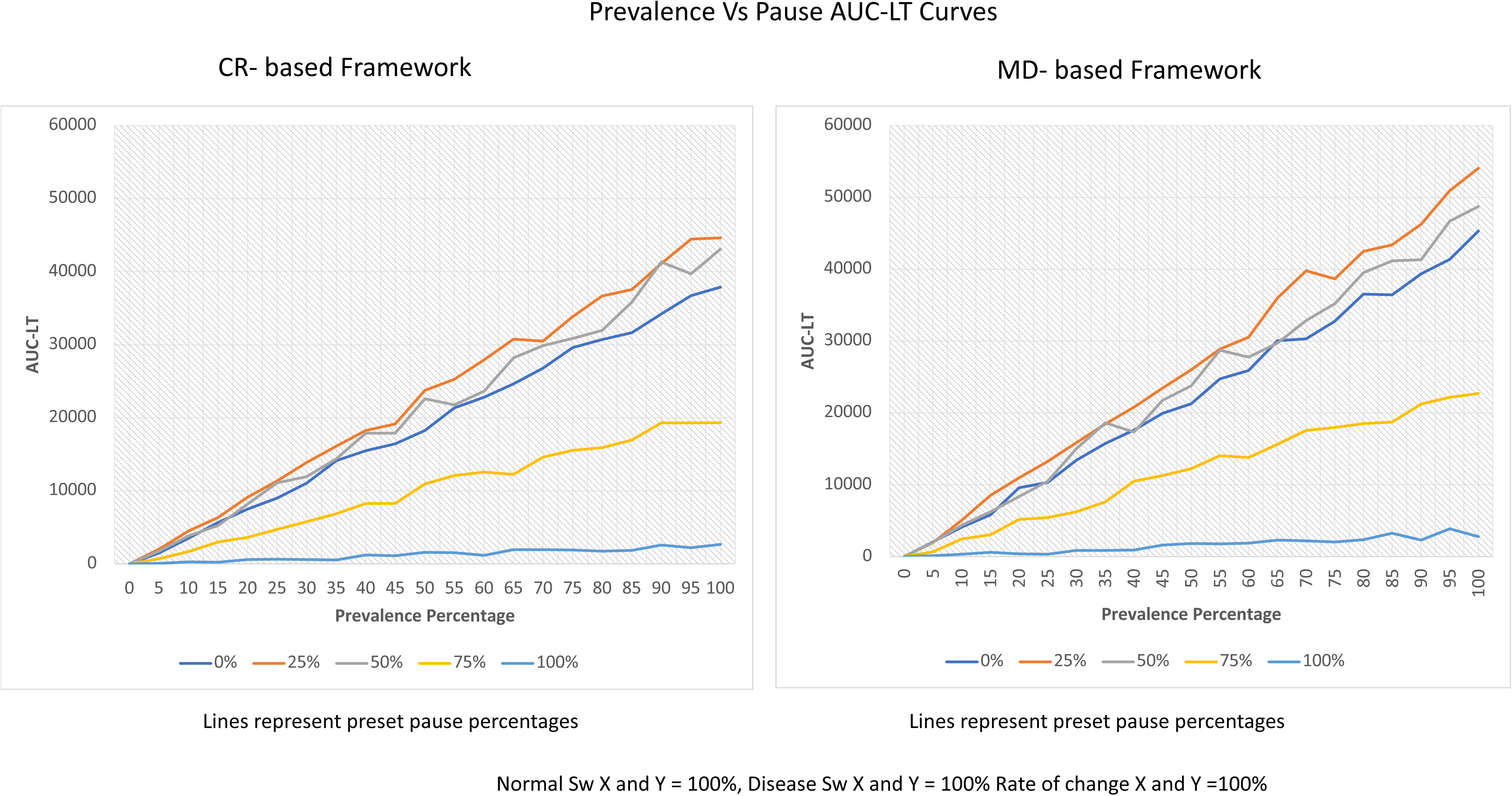
Performance Gradient Curves across Prevalence and Pause levels. These line plots represent the AUC-LT (y-axis) across a range of disease prevalence values (x-axis), with separate curves corresponding to fixed pause levels (0%, 25%, 50%, 75%, and 100%). (Left panel: CR-based framework; right: Mahalanobis Distance (MD) framework).

### 3. Variable Selection and Biological Logic

At an intuitive level, this framework depends on biologically interpretable axes to identify early consistent drift to detect subthreshold disease. To keep the intuition intact, but create a geometrically balanced behavior we chose the following methodology:

a. In the bivariate (2D) model, the chosen variables x and y moved in opposite directionality, same cutoff, and swapped normal and disease means. The details are in **Table 1**.
b. For the trivariate model, the variable z had a smaller scale and a directionality reverse to the pathology. The difference in scale was intentional to simulate real-world complexity.
c. This pattern was chosen as it is more intuitive and hence scalable when we are extending this platform to real-world ranges in future iterations.

**TABLE 1:** VARIABLE CHARACTERISTICS FOR THE IN-SILICO STUDY.

|  | DISEASE NAME | MODEL |  |  |
| --- | --- | --- | --- | --- |
| **1** | Variable | X | Y | Z |
| **2** | Name | x | y | z |
| **3** | Direction compared to pathology<br>(Increase +1, decrease -1) | 1.00 | **-1.00** | **1.00** |
| **4** | Threshold value for cutoff | 125 | 125 | 7.5 |
| **5** | Normal (stable) cases Mean | 100.00 | 150 | 6 |
| **6** | Normal (stable) cases SD | 5 | 5 | 0.3 |
| **7** | Normal (stable) cases<br>Intrameasurement standard deviation<br>(Sw- normal) | 5.00 | 5 | 0.2 |
| **8** | Early Disease Cases Mean | 150.00 | 100.00 | 9.00 |
| **9** | Early Disease Cases Standard<br>Deviation | 10 | 10.00 | 0.30 |
| **10** | Early Disease cases Intrameasurement<br>standard deviation (Sw- normal) | 5.00 | 5.00 | 0.20 |
| **11** | Early disease Mean Rate of change<br>per follow-up duration (6m) (absolute<br>value, no sign) | 5.00 | 5 | 0.2 |
| **12** | Early disease SD Rate of change per<br>follow-up duration (6m) | 2.5 | 2.5 | 0.1 |
| **14** | Expected rate of progression in<br>subthreshold cases compared to Early<br>disease (percent) | 100.00 | 100.00 | 100.00 |

### 4. The adjustments in MNR and CDS are more nuanced and are addressed using the two options below

a) For subject *u*, at visit *τ*, MNR derived from CR was calculated as

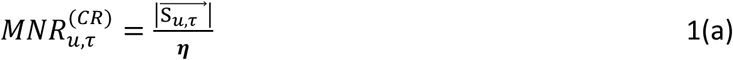
Where:
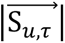 is the magnitude of the subject vector 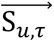
When,
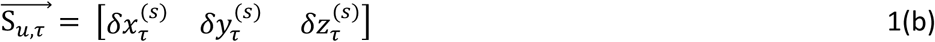
and,
*δx*^(*s*)^ *δy*^(*s*)^ *δz*^(*s*)^ are the change from baseline for variables *x*, *y*, *z* scaled by the physiological range and directionally aligned with disease progression (please see Equations 4a and 4b in Appendix A)
**b) Mahalanobis distance (MD**): MD can be interpreted as the distance of a new data point from the existing data cloud accounting for the covariance in the feature space. In MNR from CR, the intuition of dividing the noise by CR comes from the observation that 1 CR is approx. 2.77 times intrameasurement standard deviation (Sw) based on the Gaussian assumption applied to CR. However, as MD-squared follows Chi-Square distribution, the scaling factor for Sw was adjusted per the degrees of freedom (n=2 or 3) at a 95% confidence interval. Therefore, in the test subject *u*, at visit *τ*, MNR derived from Mahalanobis distance was calculated as

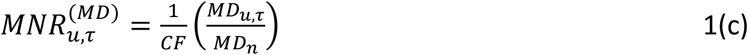
Where:

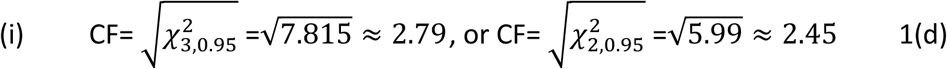
(ii) *MD_*u*,*τ*_* is the Mahalanobis distance of the subject vector 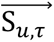 from the pooled physiological change vector *MD*_*n*_
(iii) *MD*_*n*_is calculated from a normal pool as:
For N normal subjects, with *T_j_* visits per subject
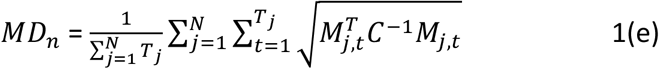
when,
(i) 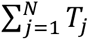 are the total follow-up visits in the normal pool.
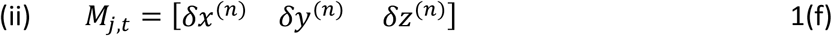
(iii) *δx*(*n*) *δy*^(*n*)^ *δz*^(*n*)^ are the change from baseline for variables *x*, *y*, *z* scaled by the physiological range and directionally aligned with disease progression (please see Equations 2a-c in Appendix A)
(iv) 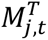 is the row vector transpose of *M_j_*_,*t*_ (i.e., 3x 1 column vector)
(v) *C*^−1^ is the inverse of the Covariance matrix for *x*, *y*, *z*
**c. Composite Drift Score:** Therefore, for subject *u*, at the visit *τ* the composite drift scores were calculated as

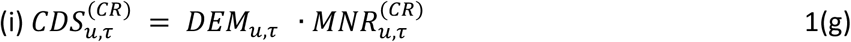
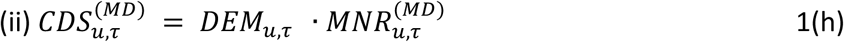
Where,

(i) *DEM*_*u*_ = cos(∅_*u*_). |cos ∅_*u*_| - please see 7b in Appendix A
(ii) ∅_*u*_is the angle between the Subject and Disease vectors - please see 7b in **Appendix A**

### 5. Comparison values for CDS and Cross-sectional parameters

**a.** CDS: A CDS of >= 1.0 was considered as an indication of a strong directional alignment with the disease vector and a drift magnitude exceeding physiological noise. Therefore, a CDS threshold of ≥1.0 was considered as the diagnostic signal of a disease-like behavior, and sustained CDS ≥1.0 over the next follow-up was considered to further strengthen this signal. By the construct of the CDS metric, the chance of an isolated CDS reading ≥1.0 in a stable normal subject is 5%, and the probability of this occurring over two consecutive follow-ups drops to 0.25%.
**b.** Threshold Cutoff: this was the value considered to be the cutoff for disease for the specific raw variable (*x*, *y or z*)
**c.** Suspect Cutoff: this was the value considered to be the cutoff for considering a suspect for the specific raw variable (*x*, *y or z*). By default, this was kept at the 80^th^ percentile between the mean of normal values and the threshold value as in 4b

### 6. Lead time calculation

To compare the drift-based system and the threshold-based system, the following time-based signals were computed:

**a. CDS Signal time *t_CDS_***

Paired CDS signals were defined as the earliest timepoint
{*τ*, *τ* + 1} ⊆ *T* such that:

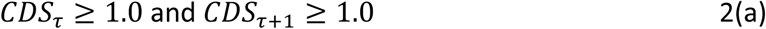
Where *T* is the ordered set of follow-up visits
The earliest such *τ* was noted as *t*_*CDS*_, representing the time point when the directional drift was first detected, hence confirmed at the next follow-up.
**b. Threshold Signal time *t_disease_***

Threshold-based diagnosis of disease state was considered when all the variables (*x*, *y* in bivariate and *x*, *y*, *z* in trivariate model) crossed the threshold for disease with a paired signal defined as the earliest timepoint(ζ, ζ + 1) ⊆ *T* such that:

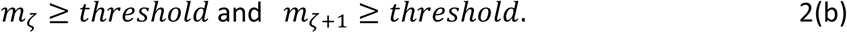
Where:
i. *m*_ζ_ is the value of the diagnostic variable*s x*, *y and z* at time ζ
ii. *T* is the ordered set of follow-up visits.
iii. *Threshold*: conventional diagnostic cutoff for variable *m*
The earliest such ζ was noted as *t*_*threshold*_, representing the first crossing of the conventional diagnostic threshold, hence confirmed at the next follow-up.
**c. Suspect Signal time *t_suspect_***

i. Threshold-based diagnosis of suspect state was considered when all the variables (*x*, *y* in bivariate and *x*, *y*, *z* in trivariate model) crossed the threshold for disease with a paired signal paired signal defined as the earliest timepoint(ζ, ζ + 1) ⊆ *T* such that:
Paired Threshold signals were defined as the earliest timepoint(ζ, ζ + 1) ⊆ *T* such that:

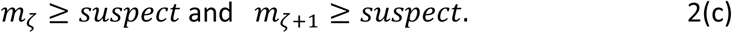
Where:

*m*_ζ_ is the value of the diagnostic variables *x*, *y or z* at time ζ
*T* is the ordered set of follow-up visits.
*Suspect* : conventional suspect cutoff for variable *m*
earliest such ζ was noted as *t*_*suspect*_, representing the first crossing of the conventional suspect cutoff, hence confirmed at the next follow-up.
**d. Bivariate and univariate worsening:**

Additionally, in the trivariate model, the following threshold crossing were also computed:
i. Bivariate suspect: when only 2 of the three variables crossed the threshold for suspect.
ii. Univariate worsening: when only 1 of the three variables crossed the threshold for suspect.
**e. Lead Time advantage *t_lead_***

To maintain comparability in the two methods, the lead time was only calculated in cases in which both CDS and either of the conventional cutoffs (suspect or threshold) were met as noted above. In those cases, the lead time was calculated as:
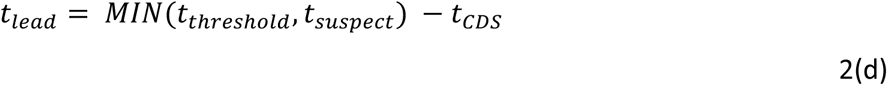
This was computed as the difference between the lower of the two values for time of paired crossing of either suspect or threshold and time for paired CDS ≥1.
**f. Area under Curve (AUC) for lead time:**

To integrate the effect of lead time and the frequency distribution of detected subjects, we define AUC-LT as
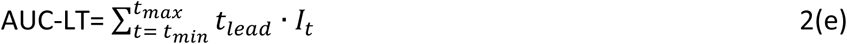
Where,
i. *t*_*lead*_ ∈

[−120,120] *months*, *range of possible lead times*(6*m x*20 *visits*)
ii. *I*_*t*_ is the number of cases with a lead time equal to t
Compared to a stand-alone metric of lead time or number of cases with CDS or conventional metrics doing better, the AUC provides a more robust analysis of the outcomes under stress simulations. For example, 300 subjects (out of 2000) with a lead time of 12 months (AUC of 3600) were considered a more robust performance for the system than 10 subjects with a lead time of 36 months (AUC of 360). This rationale becomes clearer when we stress-test the limits of the system below.

## Results

The in-silico tests were conducted using discrete steps and combinations as outlined below. The primary goals were to see the difference in AUC-LT over a range of scenarios, some mimicking biological variation and the more extreme ones simulating limits of critical failure. A clinically facing comparison is the side effects with a maximally effective dose of medication vs the maximum dose tolerable before there is irreversible toxicity. Another example from lasers is the effective usual range of depth of ablation in excimer refractive surgery and variations around that vs the dose that destines the cornea towards irreversible corneal weakness and ectasia.

We report the performance gradient curves over multiple synthetic simulations for both the CR and MD-based models. Along with this we supply screenshots of multiple dashboards with the input values and output graphs and scenarios. As noted above, these scenarios included variable disease prevalence, intermittent or fragmented progression, elevated measurement noise, elevated disease cohort noise, and accelerated parametric worsening.

The goal was to explore whether CDS, as a longitudinal index, can detect early changes even when classic diagnostic thresholds are not yet met in less-than-optimal scenarios. Both the CDS variants, one based on standard coefficient-of-repeatability (CR), and the other using Mahalanobis distance to account for multivariate correlations, were tested in two- and three-dimensional versions. Results were visualized using heatmaps and data generated from AUC-lead times offering insight into model performance under a wide range of biologically plausible and structurally plausible (even though biologically uncommon) constructs.

### Scenario specific Outcomes

1. *Base scenario (****Figure 2***): The upper two panels show CDS behavior under conditions of 100% prevalence and 0% visit-level pause, simulating a fully progressive cohort with uninterrupted (but still stochastic) directional drift. Under this setting, CDS performs very well: the Area under curve for lead time (AUC-LT) reaches ∼ 37,000 for the CR-based CDS and ∼44,000 for the Mahalanobis (MD) based CDS. The heatmaps show clear directional trend from cooler to warmer colors. In contrast, the lower two panels show the baseline scenario: 0% prevalence and 100% pause, where no subject exhibits real physiological drift. As expected, there is non-directional trend across the cohort, with the AUC being zero in both CR and MD. The accompanying heatmaps appear stochastic and non-directional, reflecting the absence of true progression and the influence of random intrameasurement noise.
2. Relationship between pause and prevalence and their effect on the AUC-LT: For all combinations for Prevalence and pause, with all other conditions being stable, the lead time was maintained for CDS based platforms compared to the threshold-based test (positive AUC-LT). This can be visualized at a more granular level in dashboard displays seen in **supplementary figures Ia, Ib and II.** For a more comprehensive understanding, we plotted the performance gradient curves for the AUC-LT values (y axis) as function of the prevalence percentage (0→100%) for 5 preset values of pause (0%,25%,50%,75%,100%), with all other conditions being stable **(Figure 3)**. The AUC-LT increased with increasing prevalence for all combinations. Interestingly higher AUC-LT values were seen in moderate level pauses (25%,50%) compared to no pause (0%) or total pause (100%).
3. Relationship between increased intrameasurement standard deviation in the normal group and AUC-LT at varying levels of prevalence and pause: *We must note that the in a normal clinical setting the intrameasurement standard deviation is well analyzed and changing it in the calculations for CDS will balance the outcomes. This simulation however is meant to analyze the effect of noisy or suboptimal baseline conditions when the normal dataset was not optimal or different from the expected cohort.* Univariate (x only) noise alteration was more forgiving than bivariate (x and y) noise alteration. As expected, there was reduction AUC noted with univariate noise. However, for bivariate noise more than 300%, the noise led to an erosion of CDS lead time and even reversal at higher values (400% and beyond) (**Supplementary figures III a-univariate, III b and III c-bivariate).**

*a.* Univariate changes gradient curves: To understand this better, we plotted performance gradient curves for AUC-LT compared to univariate change for both frameworks. First, we evaluated the outcomes (AUC-LT vs Normal Sw for X only) at 5 levels of prevalence percentage without any effect of pause (pause set at 0%) (**Figure 4a**). Therefore, this was the full progression model. After an initial reduction in AUC-LT, both the methods stabilize at around 205% (CR) to 235% (MD) to maintain a stable AUC with increasing univariate noise up to 400%. When we increased the pause rate to 50%, keeping all other simulation parameters similar, the AUC-LT increased again showing the consistent advantage of the CDS based methods in stochastic conditions over conventional threshold (**Figure 4b**). For example, for 100% prevalence at 0% pause the AUC-LT at 25% Sw -X was ∼60,000 for CR, and ∼65,000 for MD, and at ∼400% Sw-X, it was stable in a zone around 20,000 for CR and 30,000 for MD. Increasing the pause to 50% increased the AUC-LT value to ∼5000 to ∼10,000 units higher.

**Figure 4a.**
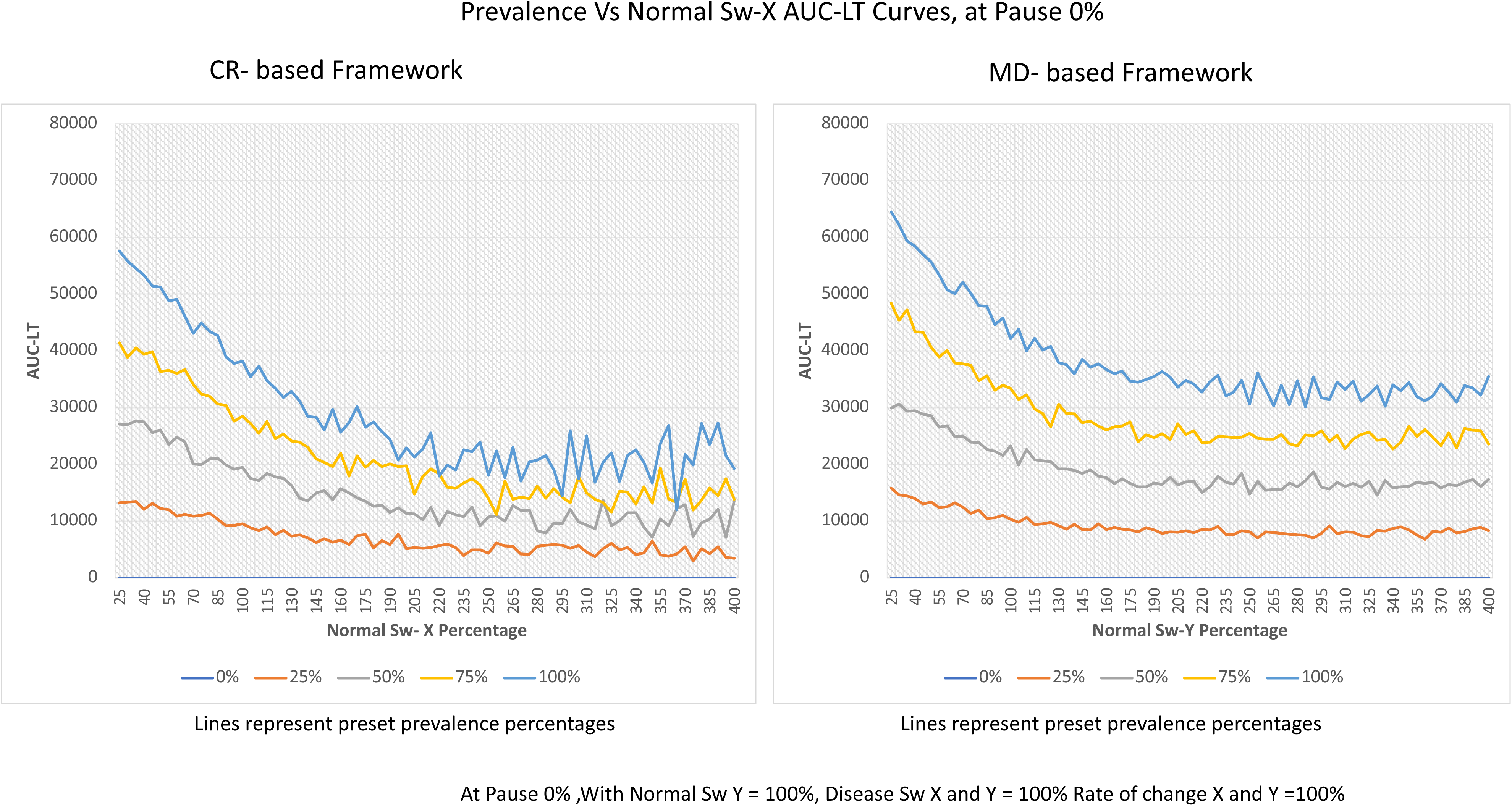
Performance Gradient Curves across Prevalence and Normal Sw-X (Intrameasurement standard deviation for one variable X for normal) at 0% pause: These line plots represent the AUC-LT (y-axis) across a range of disease Normal Sw-X values (x-axis), with separate curves corresponding to fixed prevalence levels (0%, 25%, 50%, 75%, and 100%) at aggressive progression scenario with visit level pause of 0% (all cases selected to worsen, based on prevalence percentage, progress at all visits, even though stochastically). (Left: CR-based framework; Right: MD based framework).

**Figure 4b.**
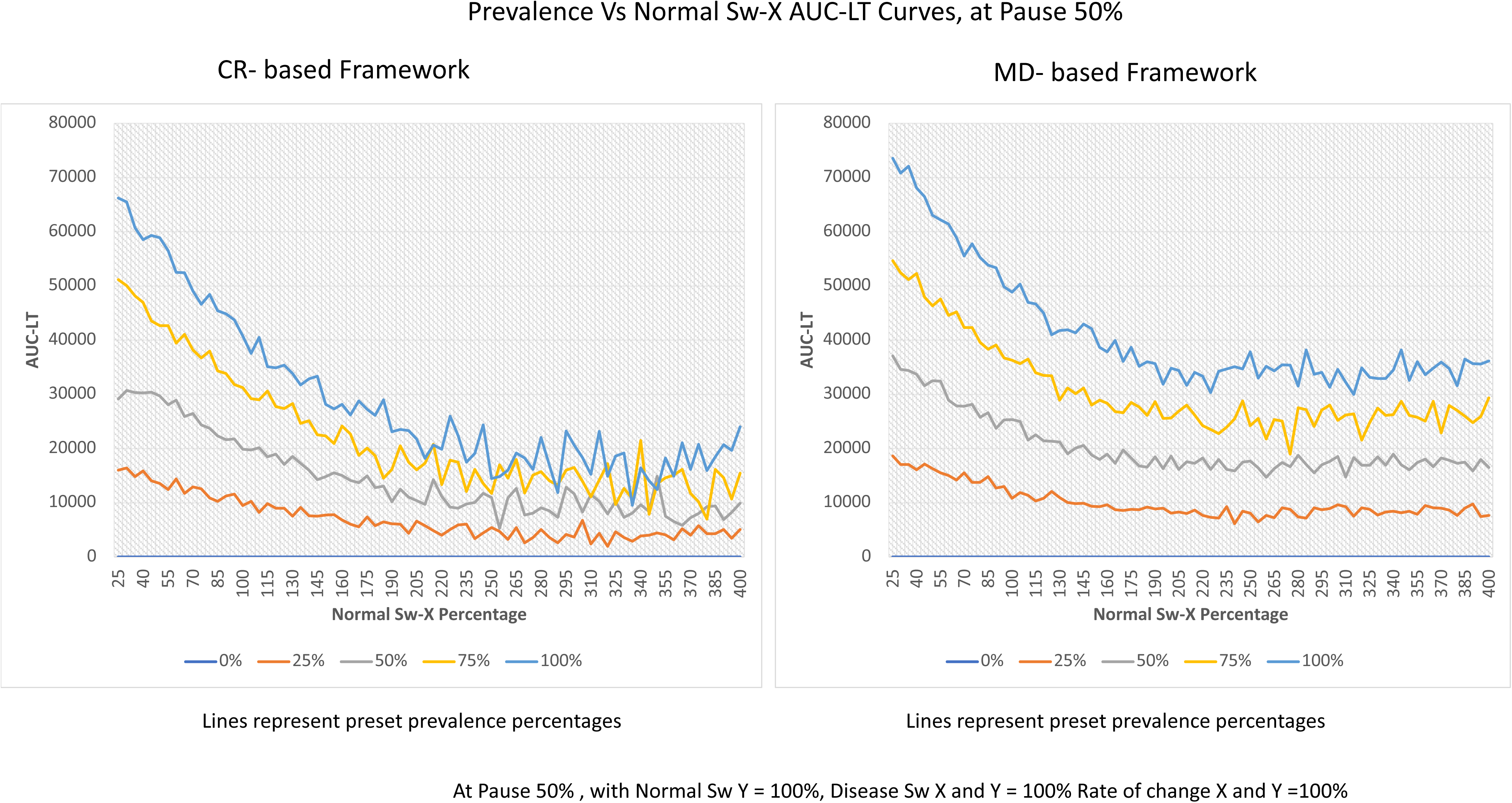
Performance Gradient Curves across Prevalence and Normal Sw-X at 50% pause: These line plots represent the AUC-LT (y-axis) across a range of disease Normal Sw-X values (x-axis), with separate curves corresponding to fixed prevalence levels (0%, 25%, 50%, 75%, and 100%) at a more biologically plausible scenario with visit level pause of 50% (for cases selected to worsen, based on prevalence percentage, only 50% visit show progression, and rest of the visits are stable) (Left: CR-based framework; Right: MD based framework).
*b.* Bivariate changes gradient curves: since we saw in the dashboards that there was a true reversal in this group compared to univariate which had stability, we analyze this group in more detail. For a set pause of 0%, the AUC-LT for bivariate Sw increase was plotted at levels of prevalence (Figure 5a). There was a consistent, near linear trend of decreasing AUC-LT with a reversal at ∼205% for CR and ∼235% for MD. In lines with the trend of better performance under more stringent test conditions, the CDS based models did better when the pause percentage was increased to 50% (Figure 5b). Even though the reversal was approximately at the same levels, the AUC stabilized at <-10,000 for increasing noise beyond the reversal point. Similar trends were seen when the prevalence was set at 50% and 100% and the pause was varied from 0 to 100% in steps of 25% (**Figure 5c and 5d**). The peak effects were seen at 25 and 50% pause as in lines with the trend seen before. This ensures verification of the trend seen in figures 5a and 5b from the vantage point of pause alteration.

**Figure 5a.**
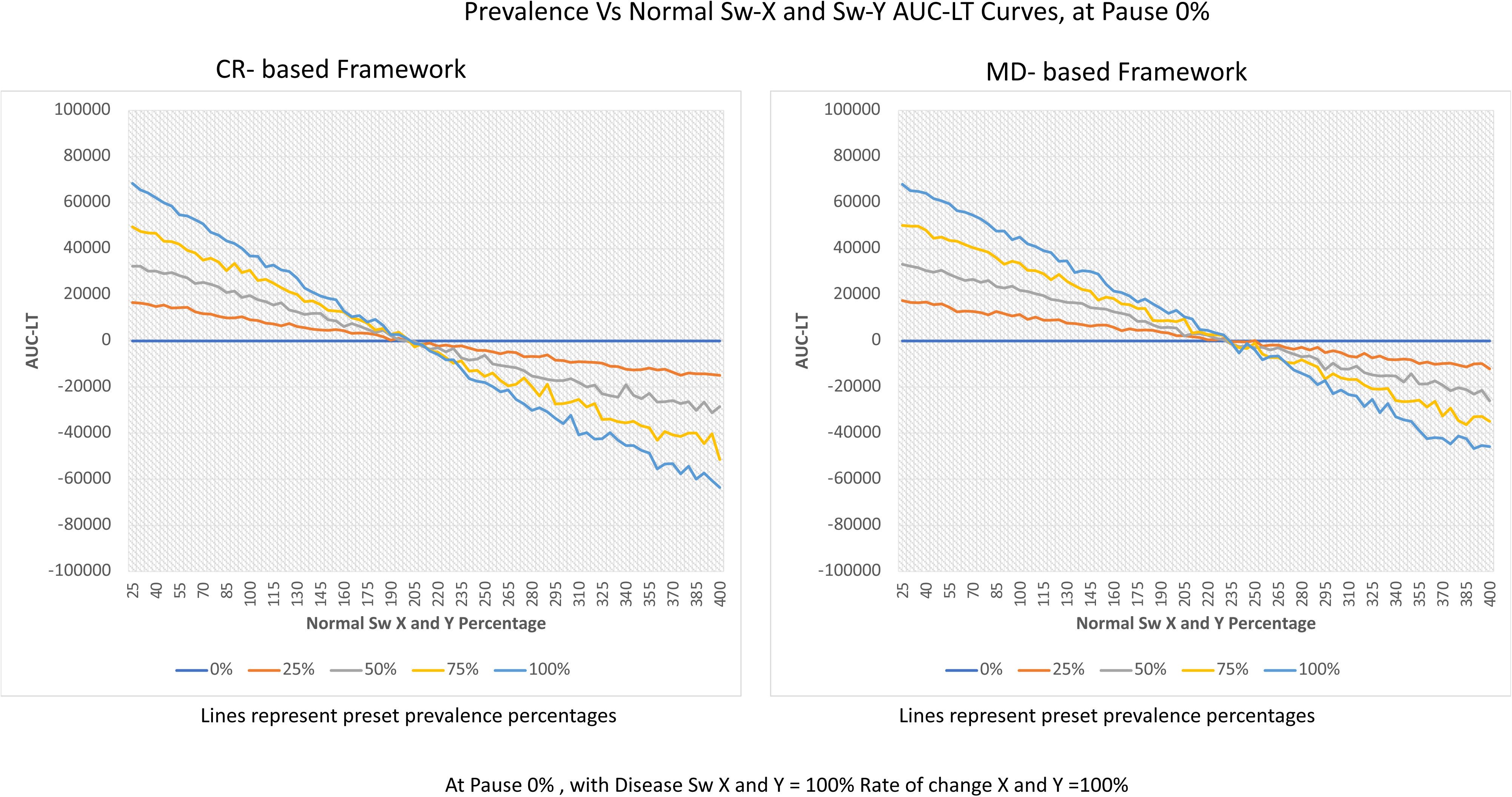
Performance Gradient Curves across Prevalence and Normal Sw-X AND Sw-Y at 0% pause: These line plots represent the AUC-LT (y-axis) across a range of disease Normal Sw-X and Sw-Y values (x-axis), with separate curves corresponding to fixed prevalence levels (0%, 25%, 50%, 75%, and 100%) at aggressive progression scenario with visit level pause of 0%. (Left: CR-based framework; Right: MD based framework).

**Figure 5b.**
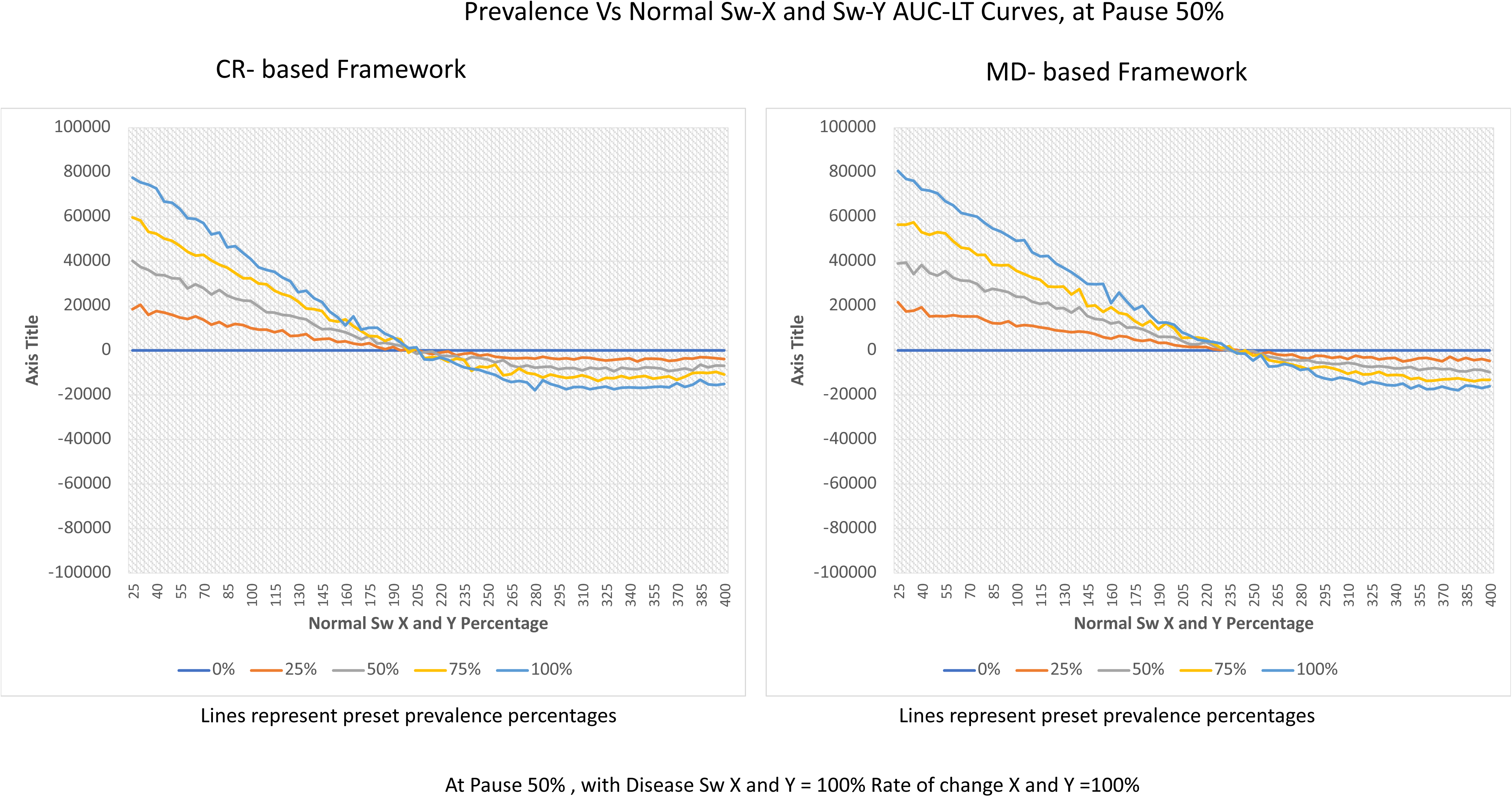
Performance Gradient Curves across Prevalence and Normal Sw-X AND Sw-Y at 50% pause: These line plots represent the AUC-LT (y-axis) across a range of disease Normal Sw-X and Sw-Y values (x-axis), with separate curves corresponding to fixed prevalence levels (0%, 25%, 50%, 75%, and 100%) at a more biologically plausible scenario with visit level pause of 50%. (Left: CR-based framework; Right: MD based framework).

**Figure 5c.**
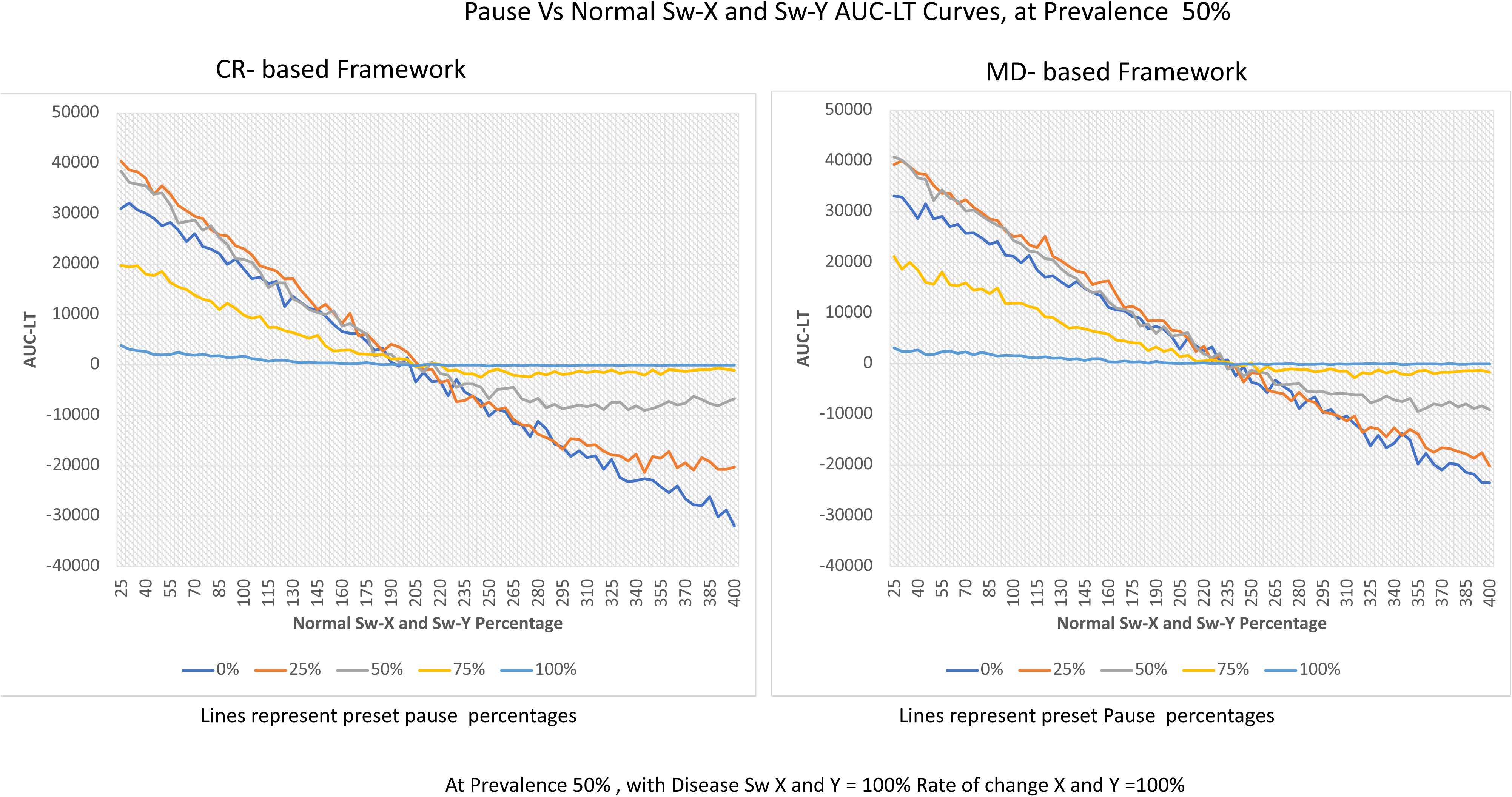
Performance Gradient Curves across Pause and Normal Sw-X and Sw-Y at 100% prevalence: These line plots represent the AUC-LT (y-axis) across a range of intrameasurement standard deviation (Sw) for both variables X and Y (x-axis), with separate curves corresponding to fixed pause levels (0%, 25%, 50%, 75%, and 100%) set at the scenario of 100% prevalence. (Left: CR-based framework; Right: MD based framework).

**Figure 5d.**
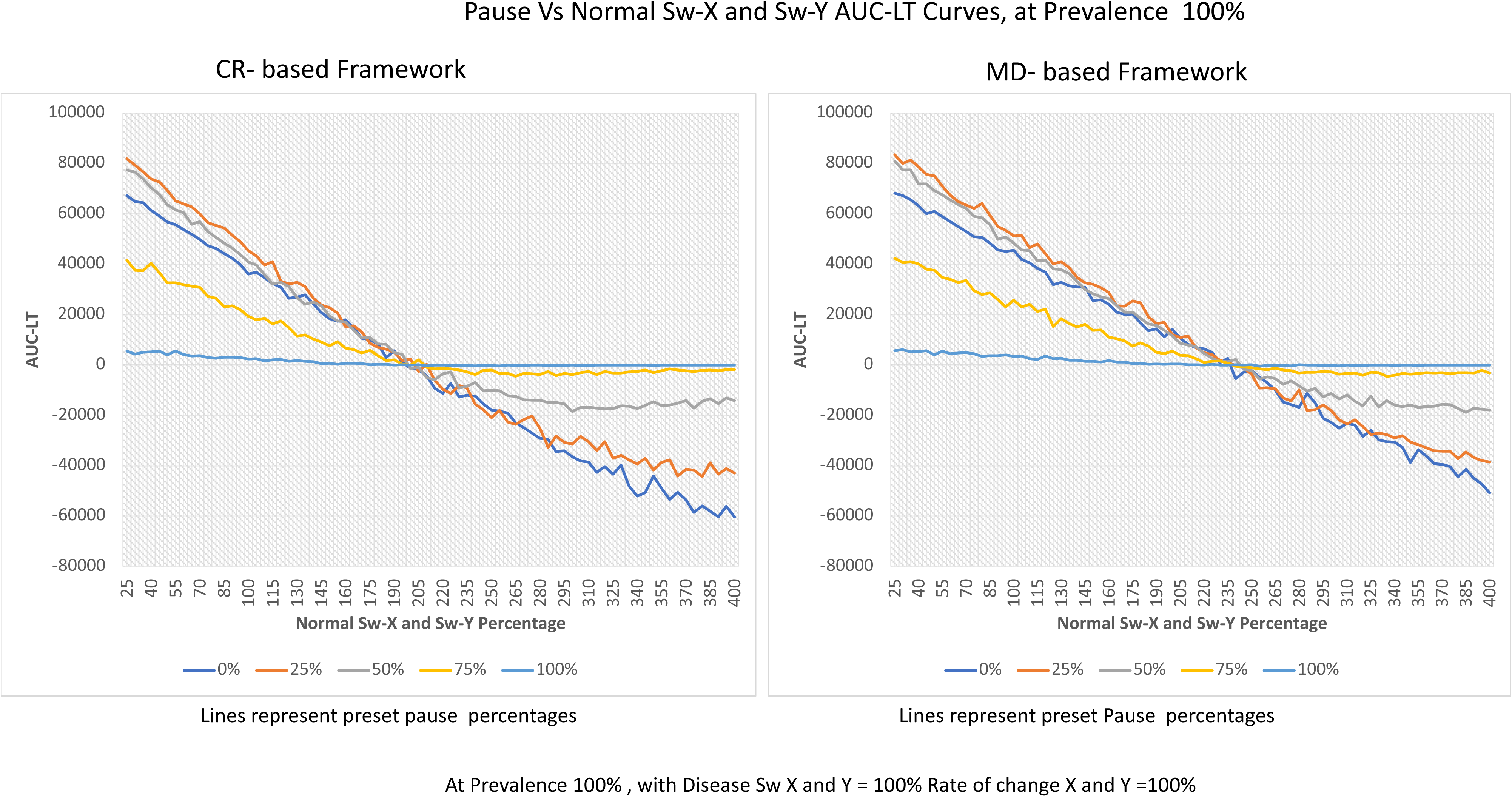
Performance Gradient Curves across Pause and Normal Sw-X and Sw-Y at 50% prevalence: These line plots represent the AUC-LT (y-axis) across a range of intrameasurement standard deviation (Sw) for both variables X and Y (x-axis), with separate curves corresponding to fixed pause levels (0%, 25%, 50%, 75%, and 100%) set at the scenario of 50% prevalence. (Left: CR-based framework; Right: MD based framework).
4. Relationship between Intra-Subject Measurement Noise and AUC-LT: The framework was also tested under conditions where noise was introduced within the group being tested itself. As expected, higher noise levels reduced signal clarity (**Supplementary Figure IV)**. Like the previous simulations, 4 different combinations (pause 0%, pause 50%, prevalence 50% and prevalence 100%) were used to evaluate AUC-LT over test subjects Sw-X and Sw-Y at varying preset prevalence and pause levels (**Figures 6 a-d).** At 0% pause, there seemed to be a smooth local maxima in AUC at around 170-200% for both frameworks (Figure 6a). For 50% pause, a trend towards stabilization was seen after 250% for both frameworks (**Figure 6b**). As expected, a combination of these trends (local maxima and stabilization) was seen when prevalence was set and pause was varied (**Figures 6c, 6d).** In terms of linear trends, the analysis of these variations could be useful in parameters that are more controlled by autoregulation (serum levels) vs structural measurements (such as bone density or corneal thickness). Even though the CDS model is meant for early sub-threshold disease, where this level of measurement variation is not expected, this separate assessment was performed to see if future iterations of the model can work in advanced but noisy data over short ranges. Noise-Only Condition: A zero-disease progression combined (100% pause) with increasing intra-subject noise led to a slow increase in AUC-LT (figures 6c and 6d). These cases likely represent coincidental directional alignment over follow-ups. Cases with values near the threshold and few random right-sided values can cause this trigger in a large dataset. Due to the random nature of noise, it is expected that over follow-up such cases will not have an increasing CDS score. However, this is an important feature for us to flag. CDS reset to a new baseline if such cases will be useful. It remains to be seen how significant this will be for us in clinical settings, however, we do expect that the second layer of the rate of change of CDS can be used to counter this in future iterations. This confirms that the model has inherent resistance to random noise and does not generate a signal in the absence of true physiological drift.

**Figure 6a.**
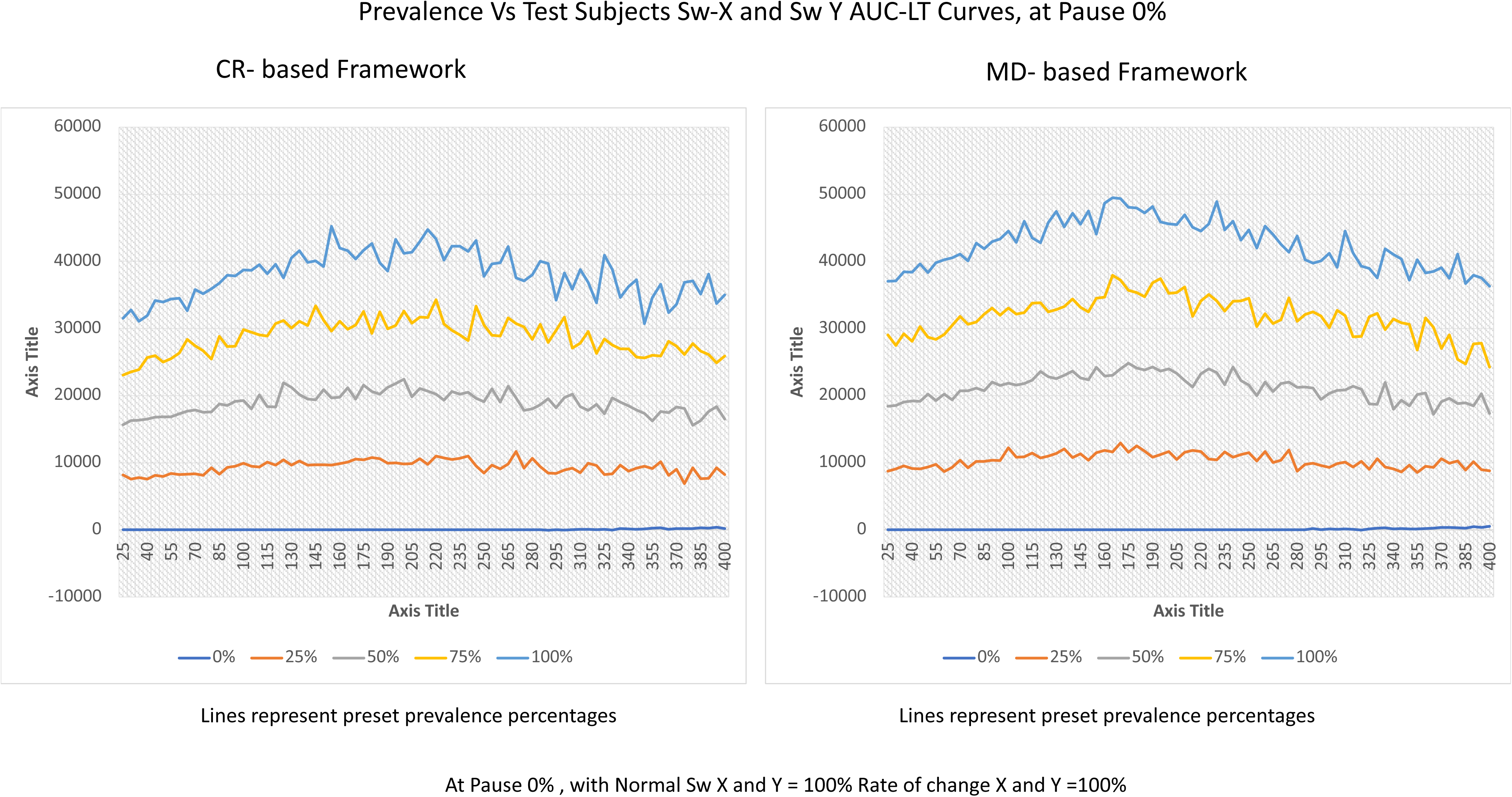
Performance Gradient Curves across Prevalence and Test Subjects Sw-X AND Sw-Y at 0% pause: These line plots represent the AUC-LT (y-axis) across a range of disease Normal Sw-X and Sw-Y values (x-axis), with separate curves corresponding to fixed prevalence levels (0%, 25%, 50%, 75%, and 100%) at an aggressive progression scenario with visit level pause of 50%. (Left: CR-based framework; Right: MD based framework).

**Figure 6b.**
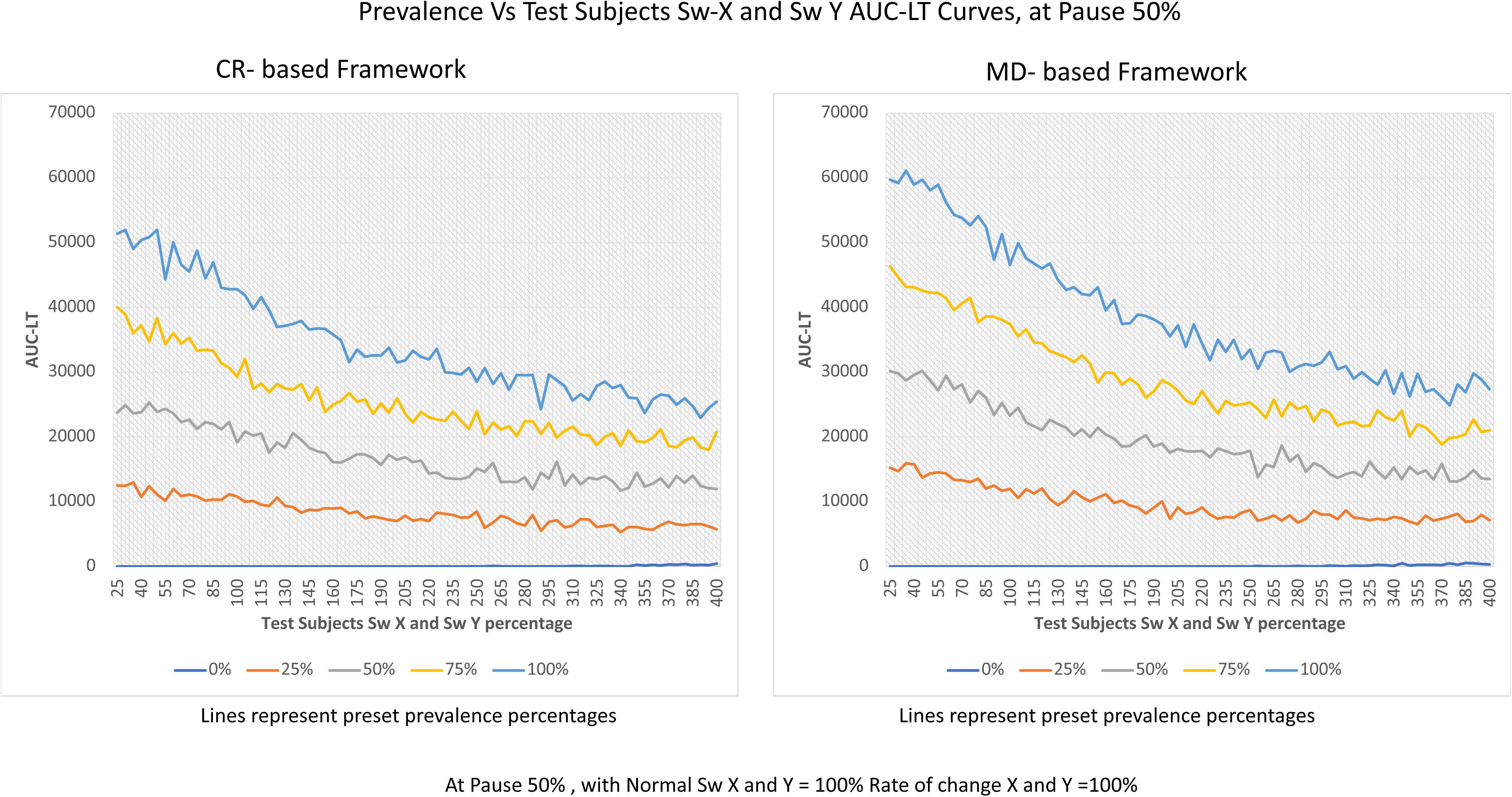
Performance Gradient Curves across Prevalence and Test Subjects Sw-X AND Sw-Y at 50% pause: These line plots represent the AUC-LT (y-axis) across a range of disease Normal Sw-X and Sw-Y values (x-axis), with separate curves corresponding to fixed prevalence levels (0%, 25%, 50%, 75%, and 100%) at a more biologically plausible scenario with visit level pause of 50% (Left: CR-based framework; Right: MD based framework).

**Figure 6c.**
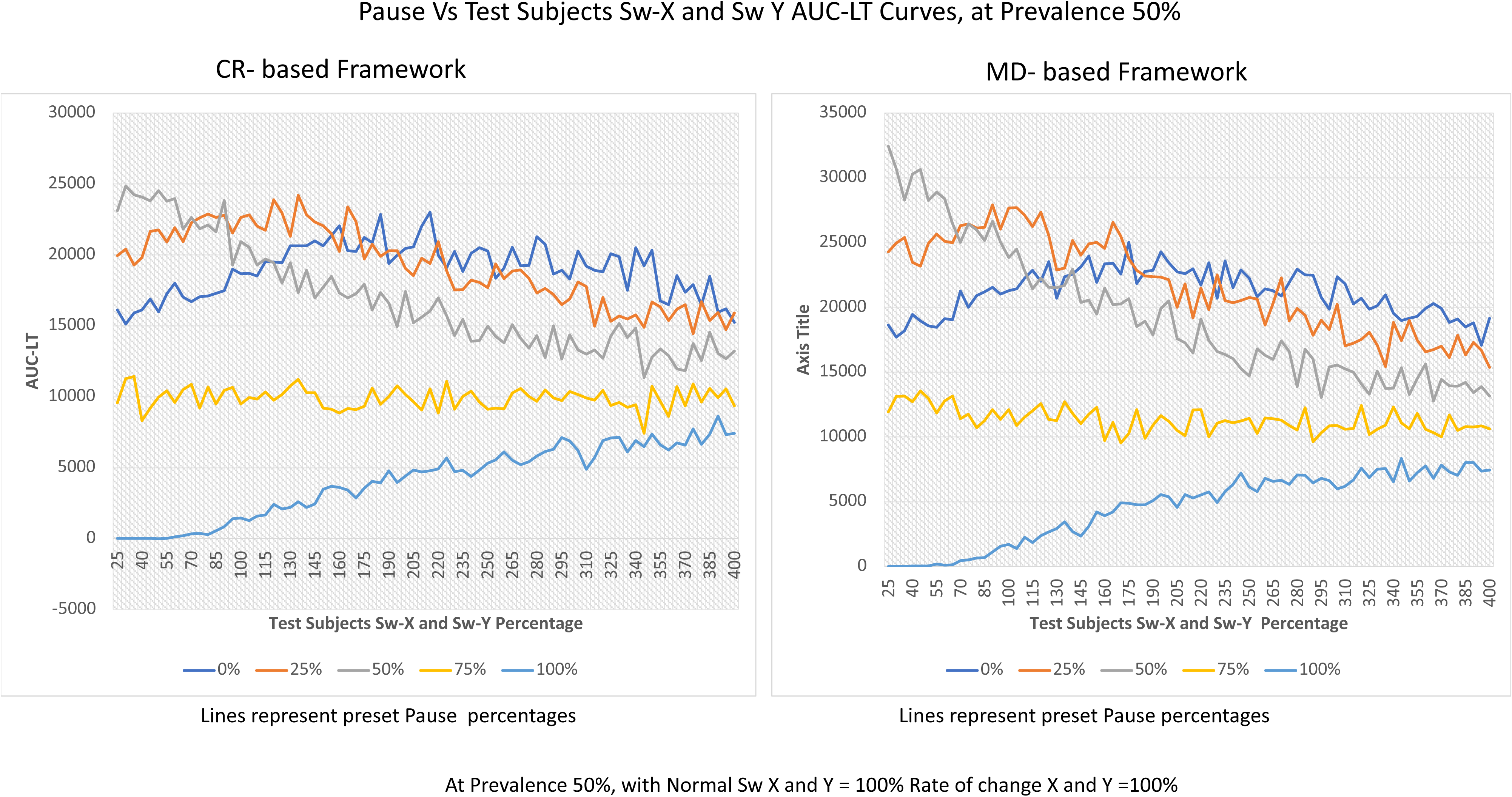
Performance Gradient Curves across Pause and Test Subjects Sw-X and Sw-Y at 100% prevalence: These line plots represent the AUC-LT (y-axis) across a range of intrameasurement standard deviation (Sw) for both variables X and Y (x-axis), with separate curves corresponding to fixed pause levels (0%, 25%, 50%, 75%, and 100%) set at the scenario of 100% prevalence. (Left: CR-based framework; Right: MD based framework).

**Figure 6d.**
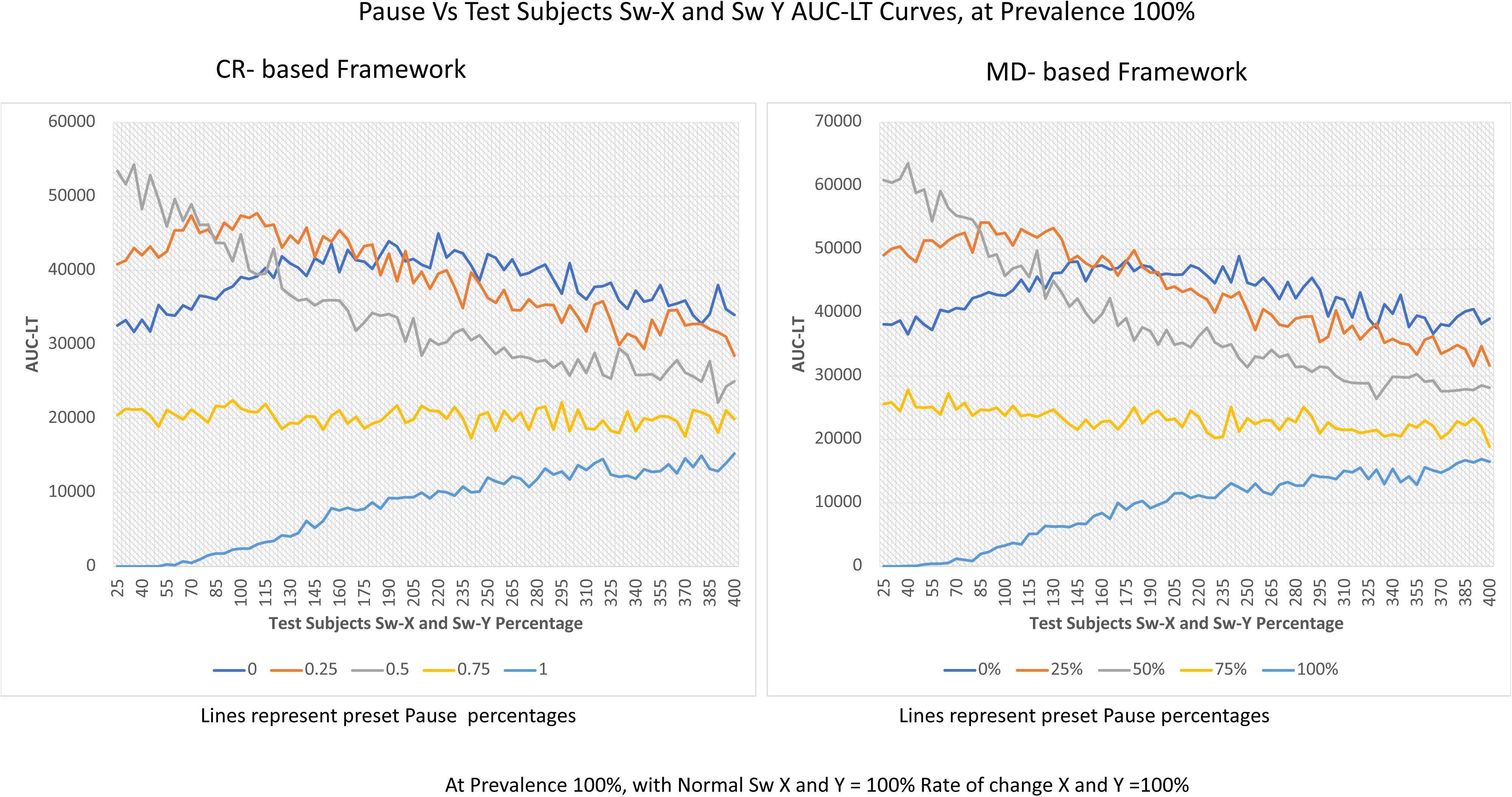
Performance Gradient Curves across Pause and Test Subjects Sw-X and Sw-Y at 50% prevalence: These line plots represent the AUC-LT (y-axis) across a range of intrameasurement standard deviation (Sw) for both variables X and Y (x-axis), with separate curves corresponding to fixed pause levels (0%, 25%, 50%, 75%, and 100%) set at the scenario of 100% prevalence. (Left: CR-based framework; Right: MD based framework).
5. **AUC-LT outcomes with combinations of varying normal and test subject noise**: Heat maps were constructed to evaluate this combination (Figures 7a, 7b). Two scenarios were tested for both frameworks at Prevalence 100%: pause 0% and pause 50%. Clear trend of higher lead time performance as seen at lower Sw values, and worse effects was seen with worsening of normal Sw. The performance of both the CR based and MD based frameworks was better with worsening Subject Sw, with MD based showing more resilience.

**Figure 7a.**
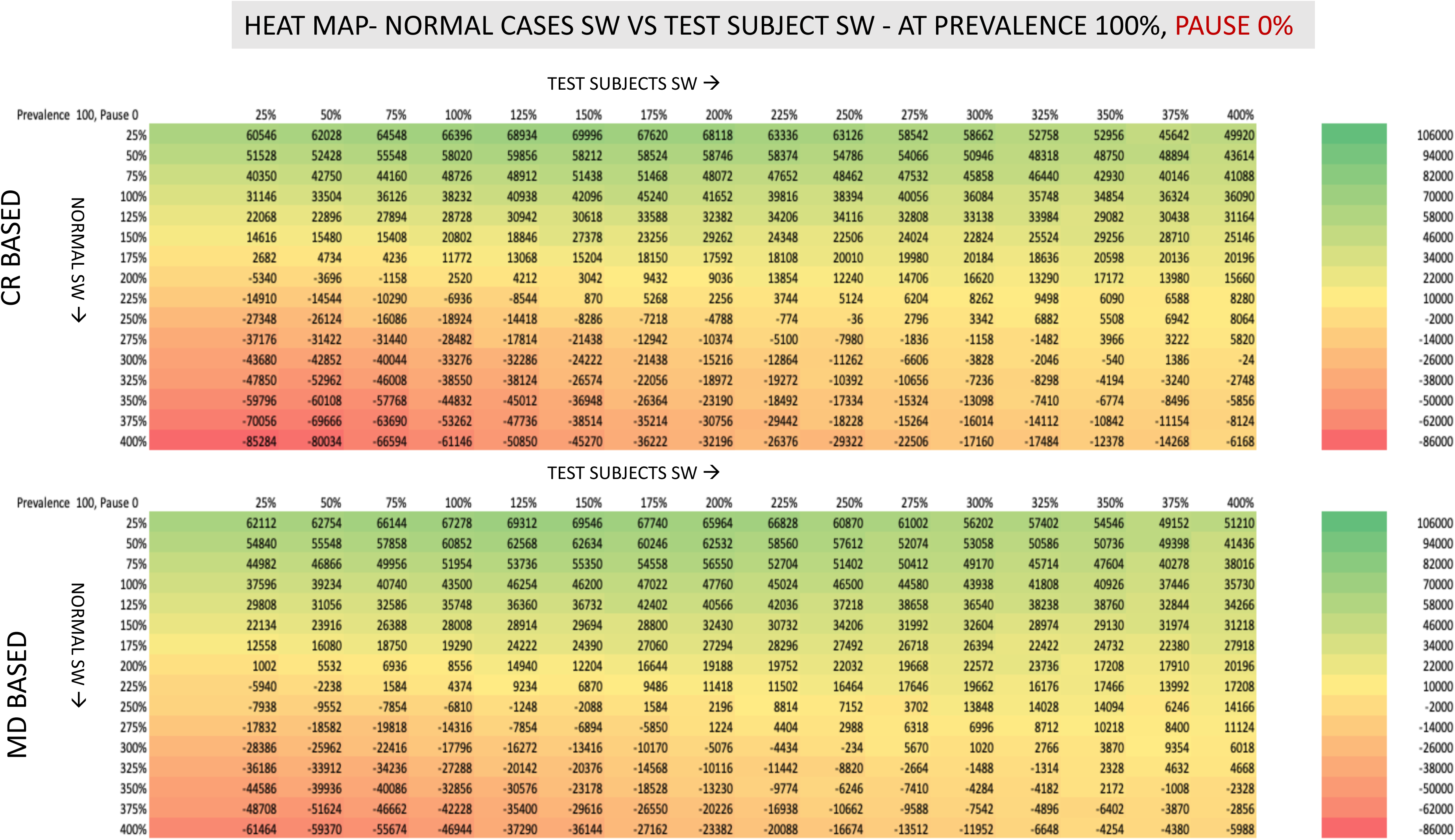
Heatmap overview of AUC-LT over a combination of test subjects Sw and Normal cases Sw: At a combination of Prevalence 100% and Pause 0%:

**Figure 7b.**
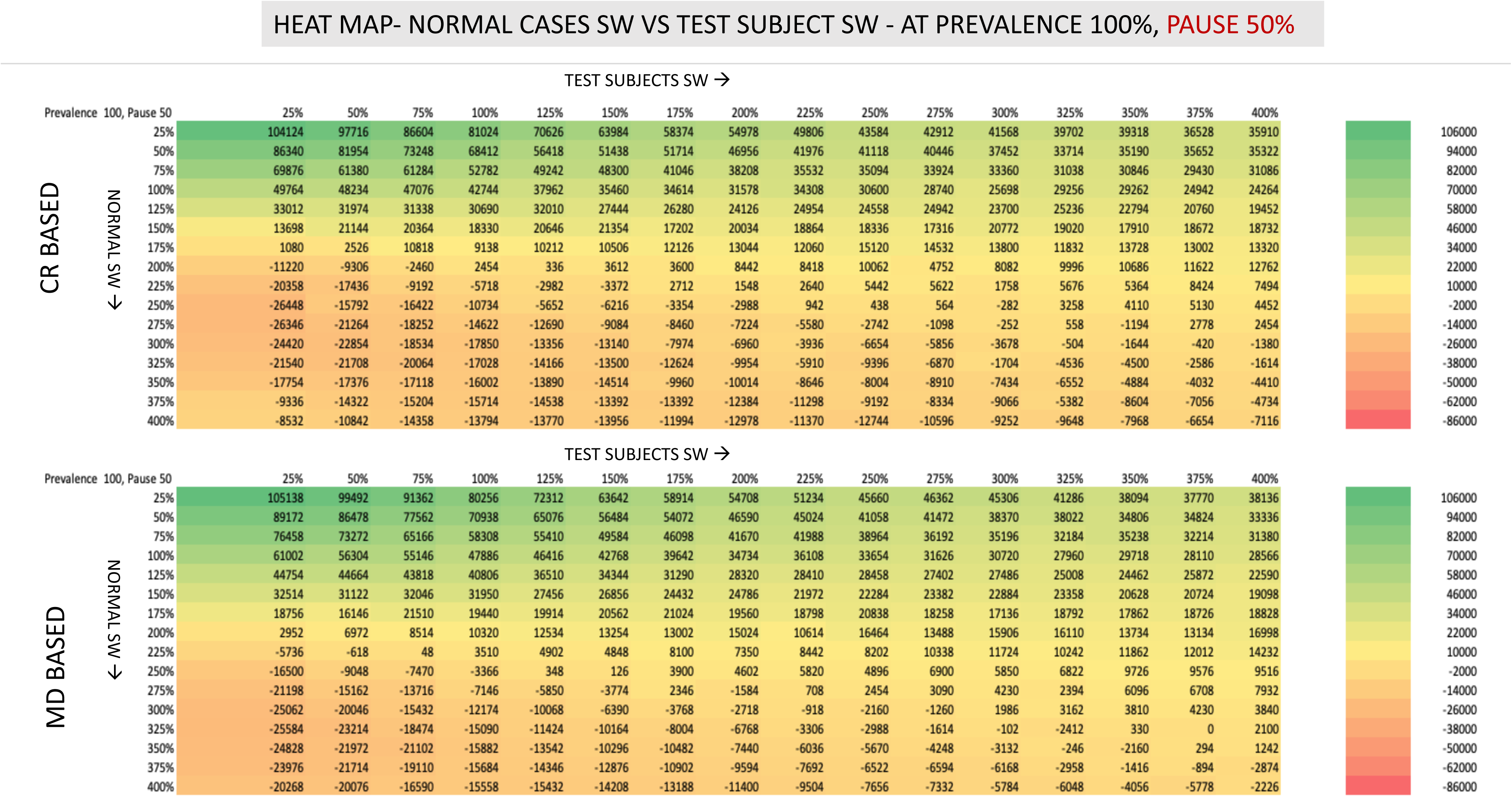
Heatmap overview of AUC-LT over a combination of test subjects Sw and Normal cases Sw: At a combination of Prevalence 100% and Pause 50%: Higher AUC-LT readings compared to no-pause scenario.
6. **Accelerated Progression:** In rapidly progressing cases, CDS was still triggered before threshold values were crossed, though lead time narrowed as the disease worsened more quickly. At extreme progression speeds (1000%), both CDS and thresholds are activated almost simultaneously (**supplementary Figure V).**
7. **Comparison between bivariate and trivariate system**: When these five combinations: Prevalence, pause, Sw normal, Sw subject and rate of change were compared for the bivariate system and trivariate system, we noted similar trends for sustained AUC-LT for both frameworks (CR and MNR). This has been represented as heat maps for easier visualization in **Figure 8a, b.**

**Figure 8a.**
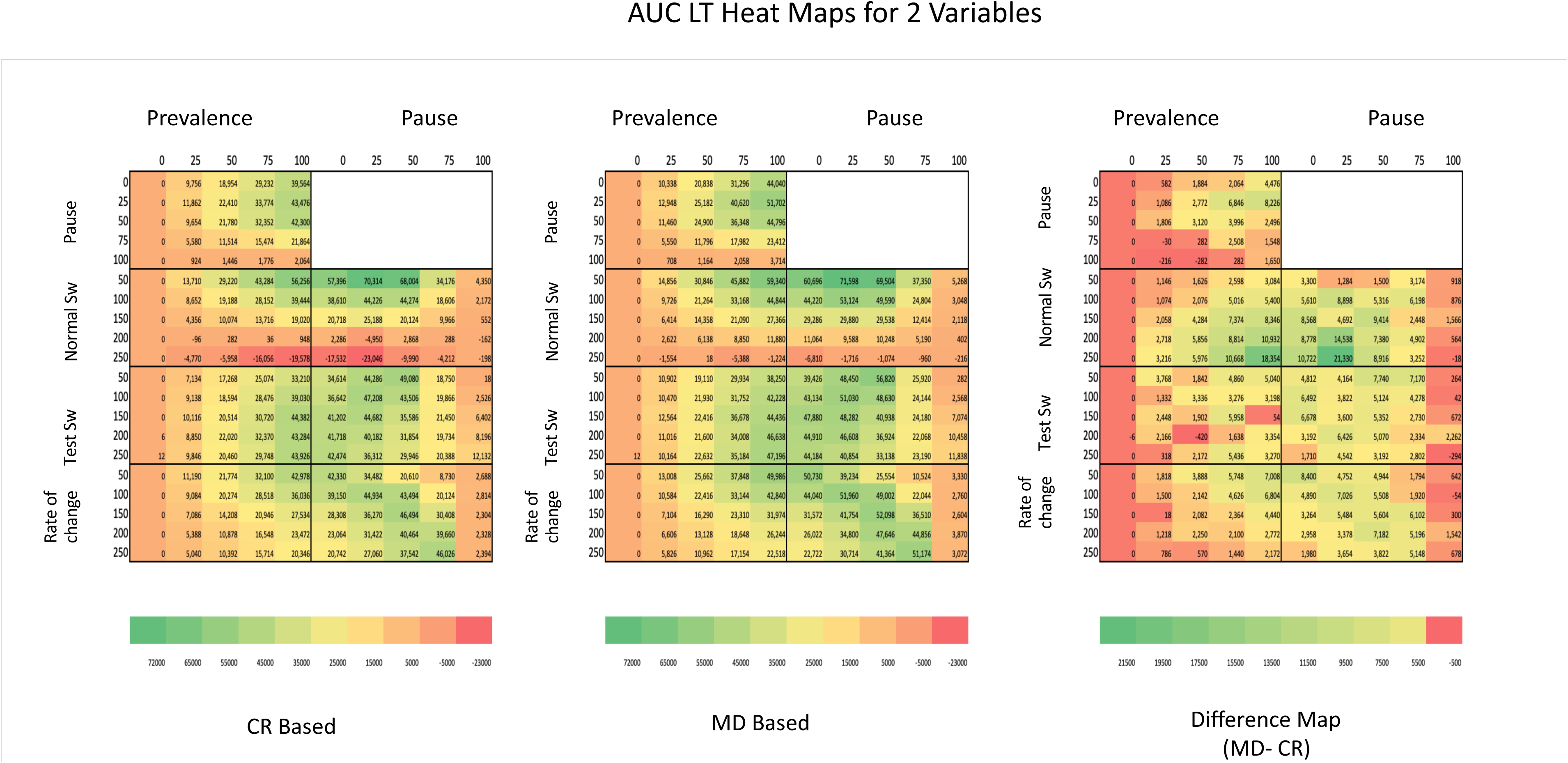
Heatmap overview of AUC-LT over a combination of parameters for the *2-variable* model, for the CR and MD based platforms and the difference maps.

**Figure 8b.**
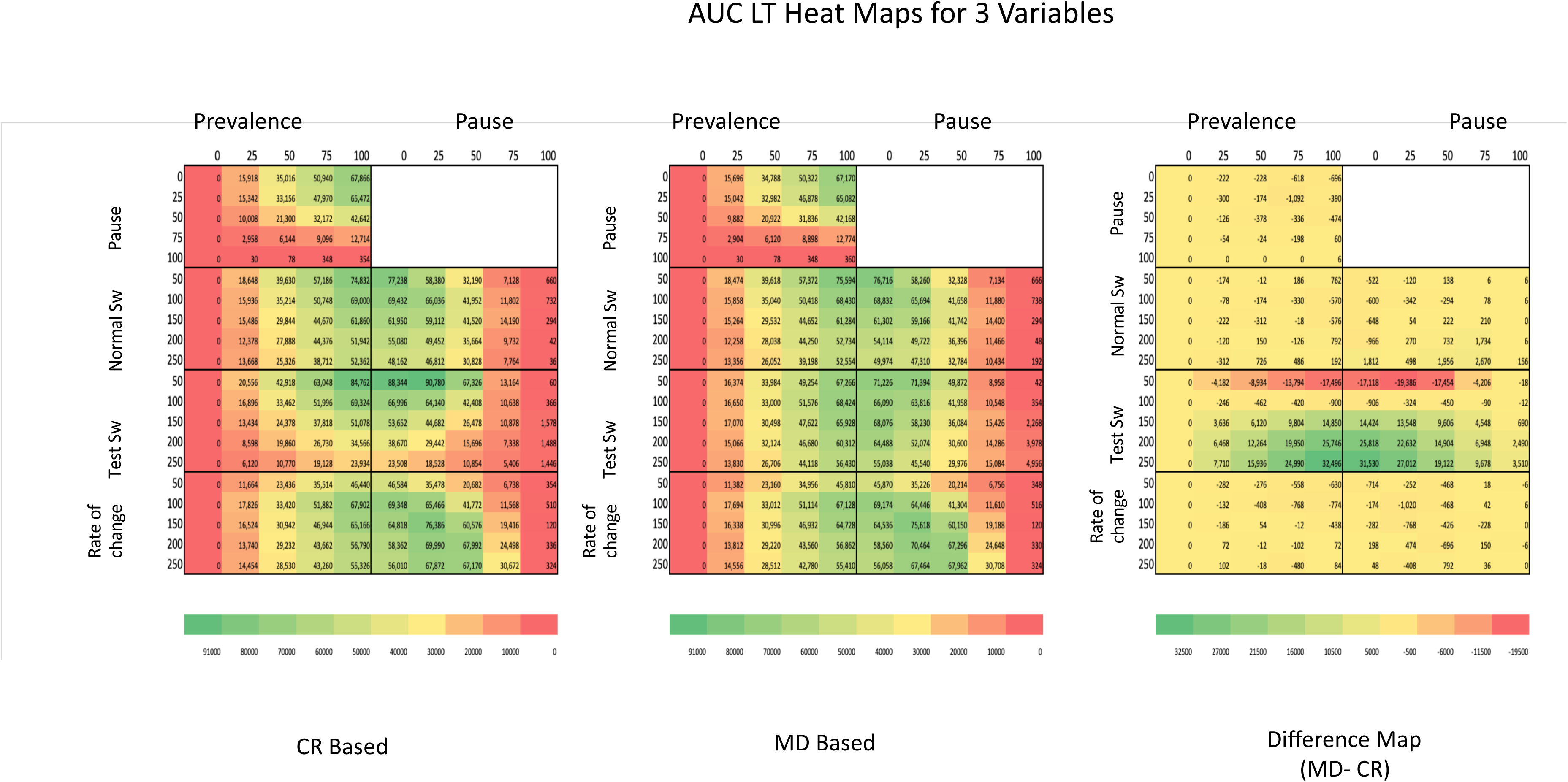
Heatmap overview of AUC-LT over a combination of parameters for the *3-variable* model, for the CR and MD based platforms and the difference maps.

## Discussion

This in-silico study was conceptualized to evaluate how the Composite Drift Score (CDS) performs under a range of stress conditions. Before we started interpreting true biological signals, this study would serve as a reference frame.

### Simulation platform and methodology

All simulations were conducted using a custom-built, modular Excel interface developed specifically for this project. This is in line with the previous study where were wanted to expand the scope of use of this platform to end-user clinicians and keep it open-sourced, editable, reproducible, and expandible for future work. We deliberately selected Excel as the initial implementation platform to strike a balance between user transparency and ease of inspection. Similar editable Excel-based frameworks have previously been used for clinical modeling and simulation, offering both pseudo-code transparency and user adaptability. For example, standardized graph-based analysis for refractive surgery outcomes is a surgeon facing Excel-based tool and has been a gold standard requirement for reporting any refractive surgery results. Similarly sizing of phakic intraocular lens for implantation is calculated on a downloadable excel model carrying the “LASSO” formula.[7] The Excel module used in our study includes hard-coded randomization blocks, adjustable scenario parameters, and conditional logic for computing CDS (both CR-based and Mahalanobis-based). Each variable and drift logic path is visible and editable, allowing full reproducibility and real-time validation of simulation behavior. This standalone spreadsheet generates Heatmaps, delta vector trends, and timepoint-specific classification outputs within the Excel environment. While we will be migrating our future implementations to the programming environment, this reference version allows for manual editability and clinician-facing interpretability during the early development phase.

We intentionally varied key parameters, disease prevalence, progression instances, measurement noise, and progression speed, to simulate both realistic and hyper-exaggerated biological variation. Our goal was to understand the expected behavior and more importantly the boundaries of this framework. This is especially relevant as the hope with this framework is to make it adaptable with modifications in different domains and settings.

### Findings from the simulations

The multiple simulation scenarios were chosen as both as continuous trends and discrete data points, and these trends can be duplicated, extrapolated, or re-iterated with a different set of variables. In this study, CDS showed predictable and clinically plausible behavior. It identified directional drift well before conventional diagnostic thresholds were crossed, particularly in cases with slower progression, early disease, or intermittent worsening. The net performance per subject (mean lead time) was relatively unaffected by prevalence compared to the traditionally held view about the prevalence of a disease affecting the positive predictive value of the conventional, threshold-based test. This is of significance if detecting trends suggestive of less prevalent diseases by longitudinal drift assessment methods such as CDS.

We also assessed the overall lead-time advantage CDS provides. In most biologically relevant scenarios, both CDS methods (CR and MD) flagged directional change a few years before conventional thresholds were met. To quantify this more robustly, we introduced a second measure, the area under the curve for lead (AUC-LT), which incorporates both the timing and frequency of early detections. AUC-LT proved useful in detecting the overall performance of the system, and an important consideration from a resource utilization perspective. In clinical settings, a broader population-level benefit may matter more than a handful of extreme outliers. So even though CDS was conceptualized to be a granular, subject-level metric, the AUC demonstrates its possible potential at a population pool level as well.

Slow and patchy progression of disease is a concern in many clinical settings. CDS also performed well in scenarios where disease progression was fragmented. In these simulations, patients experienced periods of stability interspersed with progression. Even in cases with a 50– 75% pause rate, CDS continued to identify early directional change in those who eventually progressed. At 100% pause, i.e., no actual drifts, CDS did not trigger solely based on normally seen noise. As its disease parameter has its own variation in noise and that can be randomly directional, it seems intuitive that a directionally aware measure such as the Mahalanobis distance may be most robust to noise compared to CR, especially when multiple parameters are used. This is not a fallacy with the CR style method but is rather an advantage of considering the covariance cloud, which the MD does. It remains to be seen though if this statistical benefit leads to a real-time benefit over the more intuitive, clinician-facing interpretation of CR. Interestingly, the CDS model performed better under scenarios of symmetrically high noise in both the normal and test groups than in scenarios where only one group was noisy. This stems from the construct of the metric as it depends both on the alignment (DEM) and the magnitude (MNR). Therefore, it is not simply measuring signal distortion but is also capturing the directionality of drift. When both populations are noisy, but that directionality is aligned, the system can still detect sustained deviation from physiological norms. However, when the normal group is stable and the test group becomes noisy, or vice versa, the direction of the drift becomes less consistent, or non-directional, the beneficial performance of the metric over conventional cutoff based thresholds decreases.

The decrease in lead time in highly accelerated progression is expected but there were insights into this. The mathematical behavior reflects the clinical reality that in acutely deteriorating patients, all diagnostic systems will perform. This is similar to all grading systems being able to diagnose advanced disease. Still, CDS consistently captured the onset of directional change, suggesting its utility as an early warning signal. This insight also helps in a possible exploration of this system in more sub-acute settings compared to what we initially visualized for this system (chronic progressive diseases)

### Findings from extreme scenario visualizations

These findings, where AUC from CDS becomes less than AUC from threshold, the AUC-LT turns negative. Some of the trendlines (such as that of combined Sw of normal cases X and y) show how AUC-LT drops off as a continuous function, therefore there is a clear zone of optimal performance, followed by a trend of worsening that is gradual. This is in line with most diagnostic or imaging systems which are clinically facing.

### Three-variable expansion

analysis of the three-dimensional framework suggested similar trends to the two-dimensional framework, proving that this is a scalable model for a higher dimensional analysis.

### Future trends

At this stage, our goal is not model selection or fine-tuning, but rather to demonstrate that both the CR-based and Mahalanobis-based CDS models function reliably under a range of biologically plausible stress scenarios. While the Mahalanobis variant occasionally shows increased resilience, especially under noise-heavy conditions, without granular long term clinical data, in-silico superiority is not meaningful. CR remains the more clinically established approach for managing inter-measurement variability and has long served as the practical ground truth in diagnostic modeling. Mahalanobis may serve as a comparative benchmark, but CR offers an important interpretability advantage. Until long-term clinical data become available, we believe both versions should be used with the caveats in mind.

### Summary

In summary, this stress-testing study demonstrates the structural reliability and therefore gathers further support for clinical potential of the CDS framework. It performs as intended in synthetic scenarios mimicking subthreshold and slow-progressing disease as was demonstrated in the previous study. It continues to do well in silico setting testing its biologically plausible and extreme situation limits. It began to fail when expected, under extreme noise, in a gradual, understandable, and therefore predictable way. This is useful as it guides us for its exploration in clinical settings. Future studies using will give more insights into the performance of this framework and its benefits in the real-world settings.

## Data Availability

All data produced in the present study are available upon reasonable request to the authors

## Supplementary Figures

**Supplementary Figure I.**
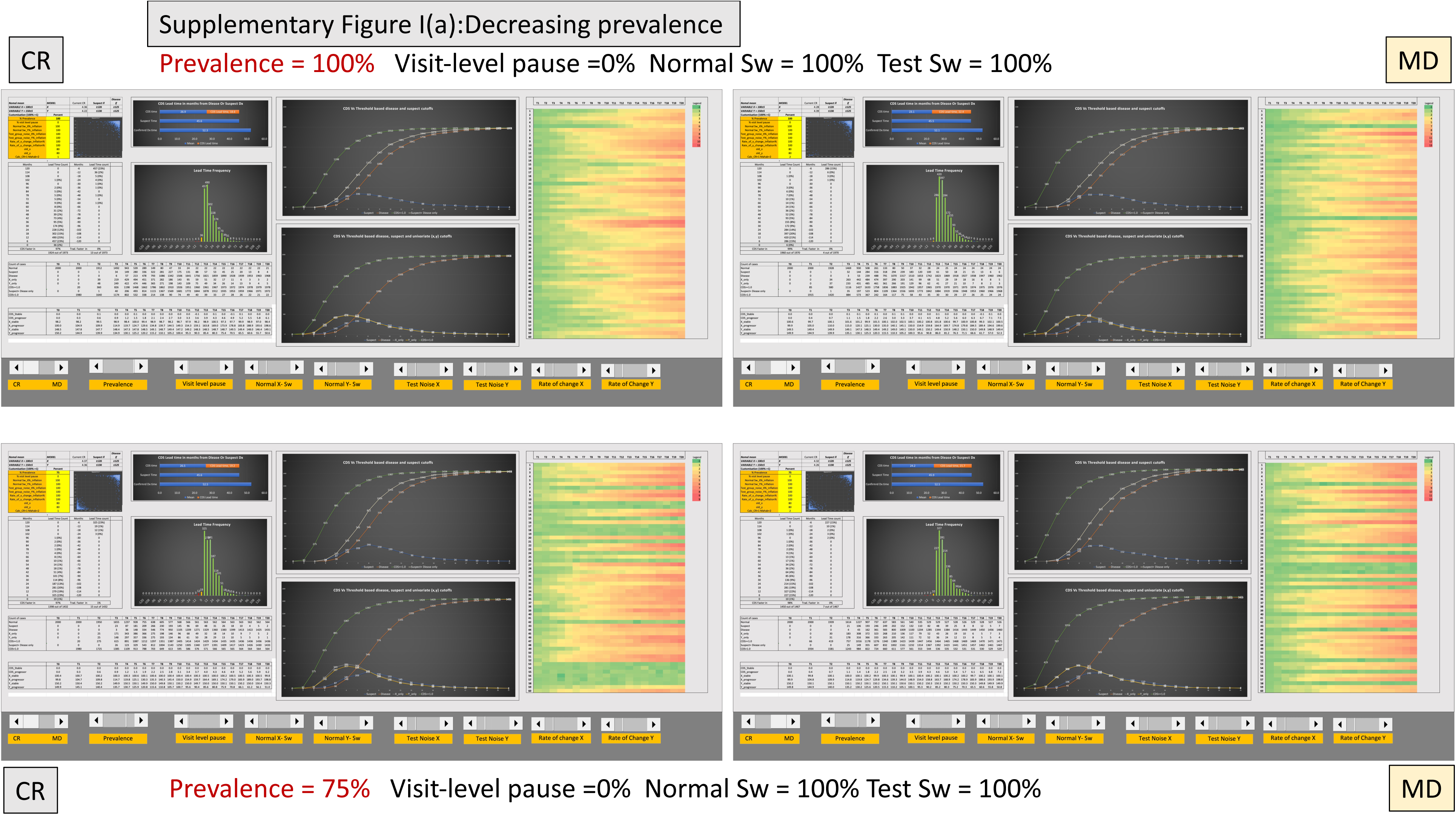
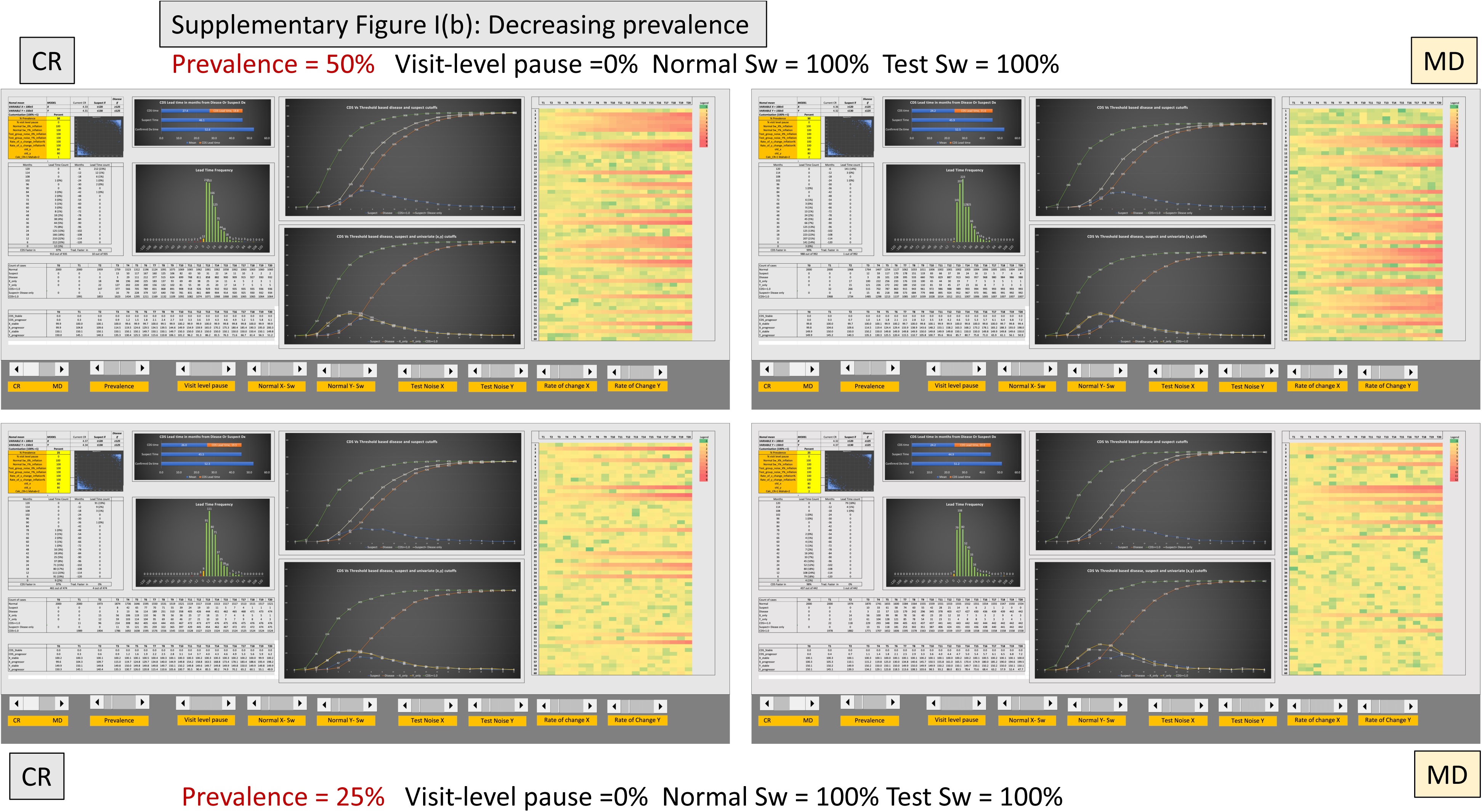
**a, b**: Effect of decreasing prevalence on framework performance. CDS signal remains robust despite decreasing disease prevalence from 100% to 25%. Both CR-based and Mahalanobis-based models maintain early directional drift detection and lead time advantage. Even at 25% prevalence, where fewer subjects progress, CDS continues to outperform thresholds by identifying progression earlier, with the Mahalanobis model showing slightly better performance.

**Supplementary Figure II.**
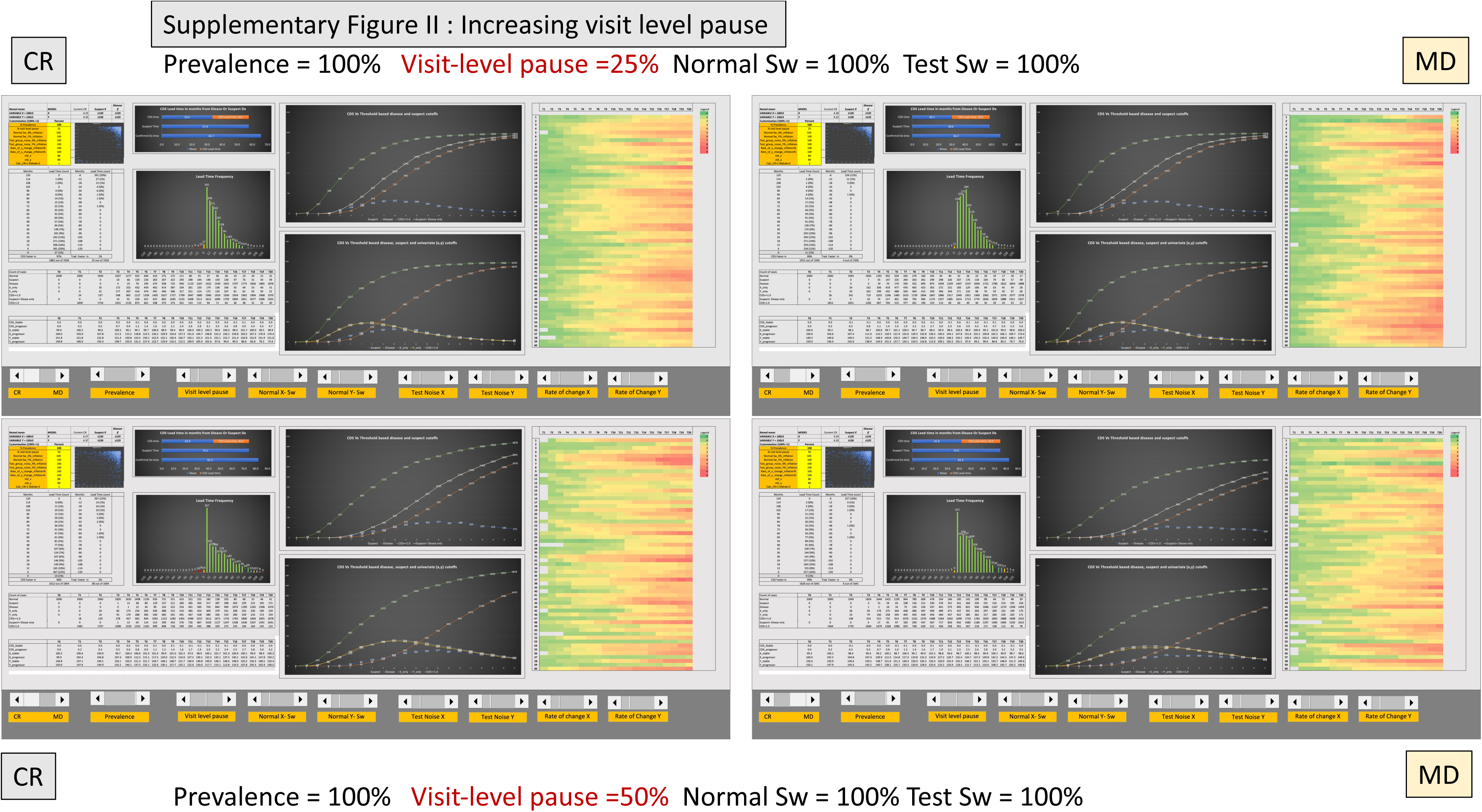
**a, b**: Effect of increasing visit level pause on the framework performance Visit-level pause introduces intermittent progression, mimicking real-world disease fluctuation. CDS maintains lead time advantage at 25% and 50% pause, especially in the Mahalanobis model. Under 50% pause with added noise, CR-based performance degrades, while Mahalanobis (MD) remains stable. This highlighting MD’s resilience to population-level measurement variability.

**Supplementary Figure III.**
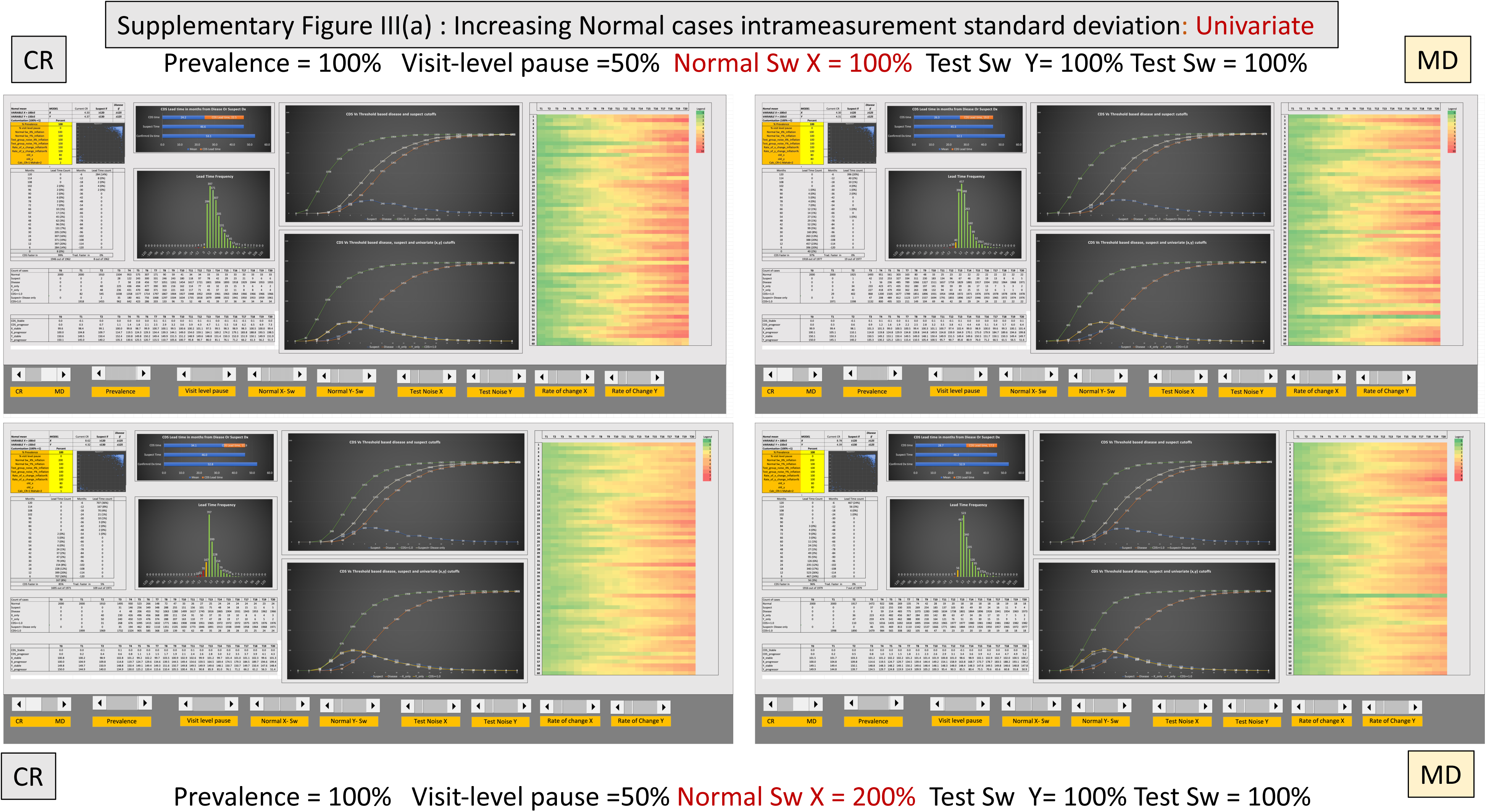
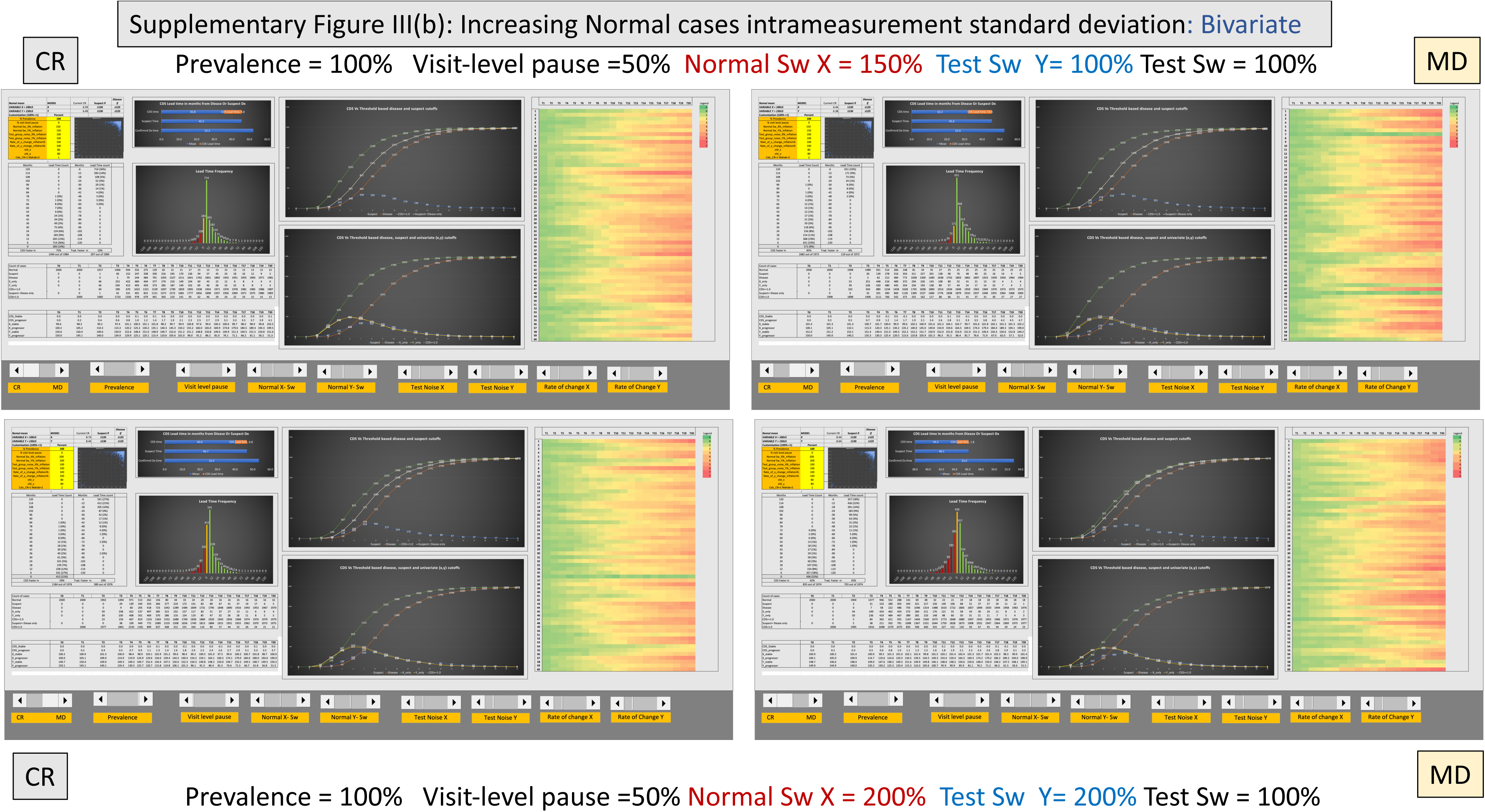
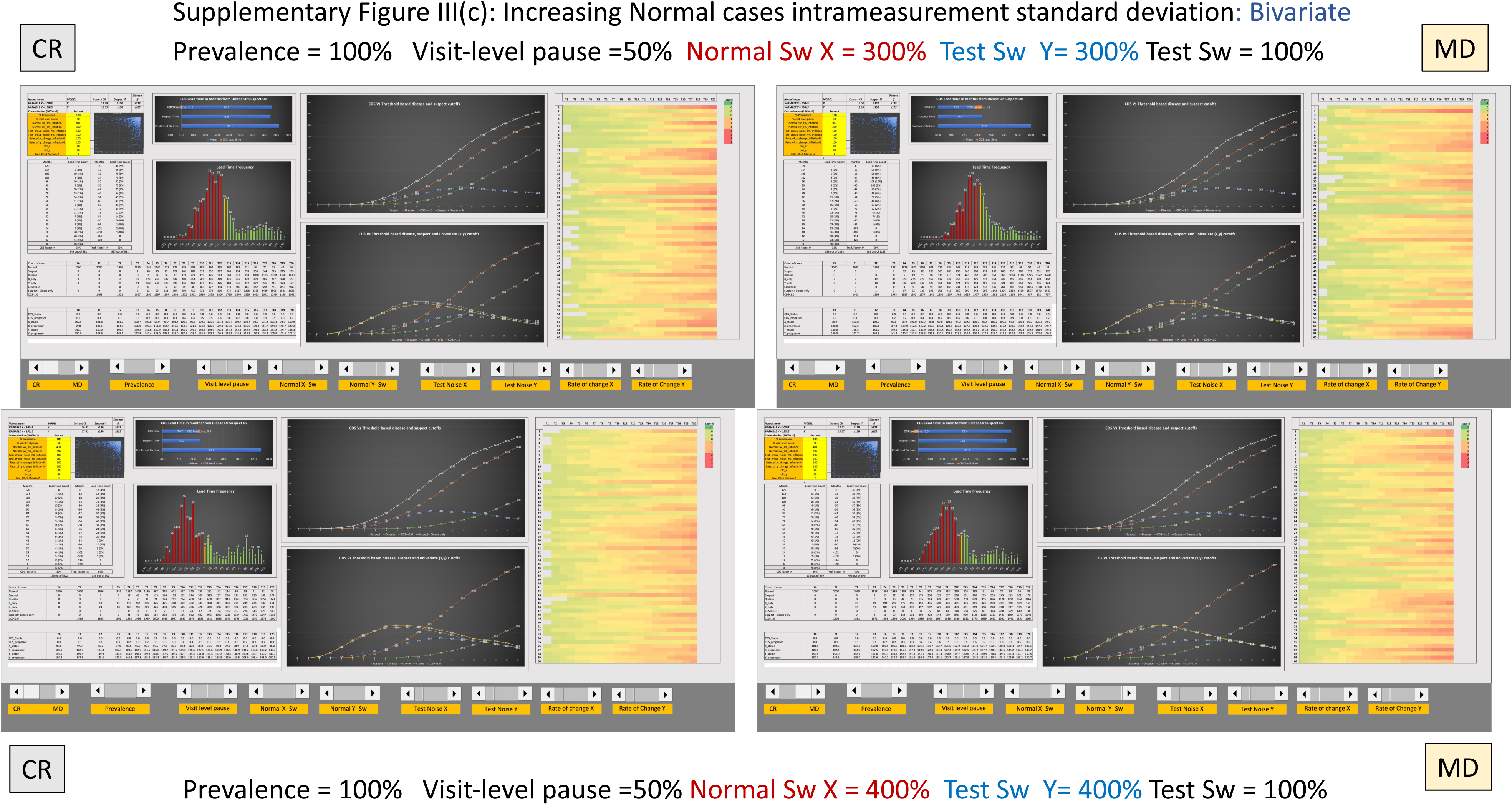
**a-c:** Effect of normal eyes intrameasurement standard deviation (Sw) (used to define ‘noise’). Univariate worsening is less strenuous on the framework (III a) compared to bivariate worsening (III b-c) Reference noise inflation reduces CDS performance as Sw increases from 150% to 400%. At 150%, both models retain acceptable drift signal, but beyond 300%, directional coherence deteriorates and the lead time benefit of CDS continues to contract, with early signs of negative AUC around 400%

**Supplementary Figure IV:**
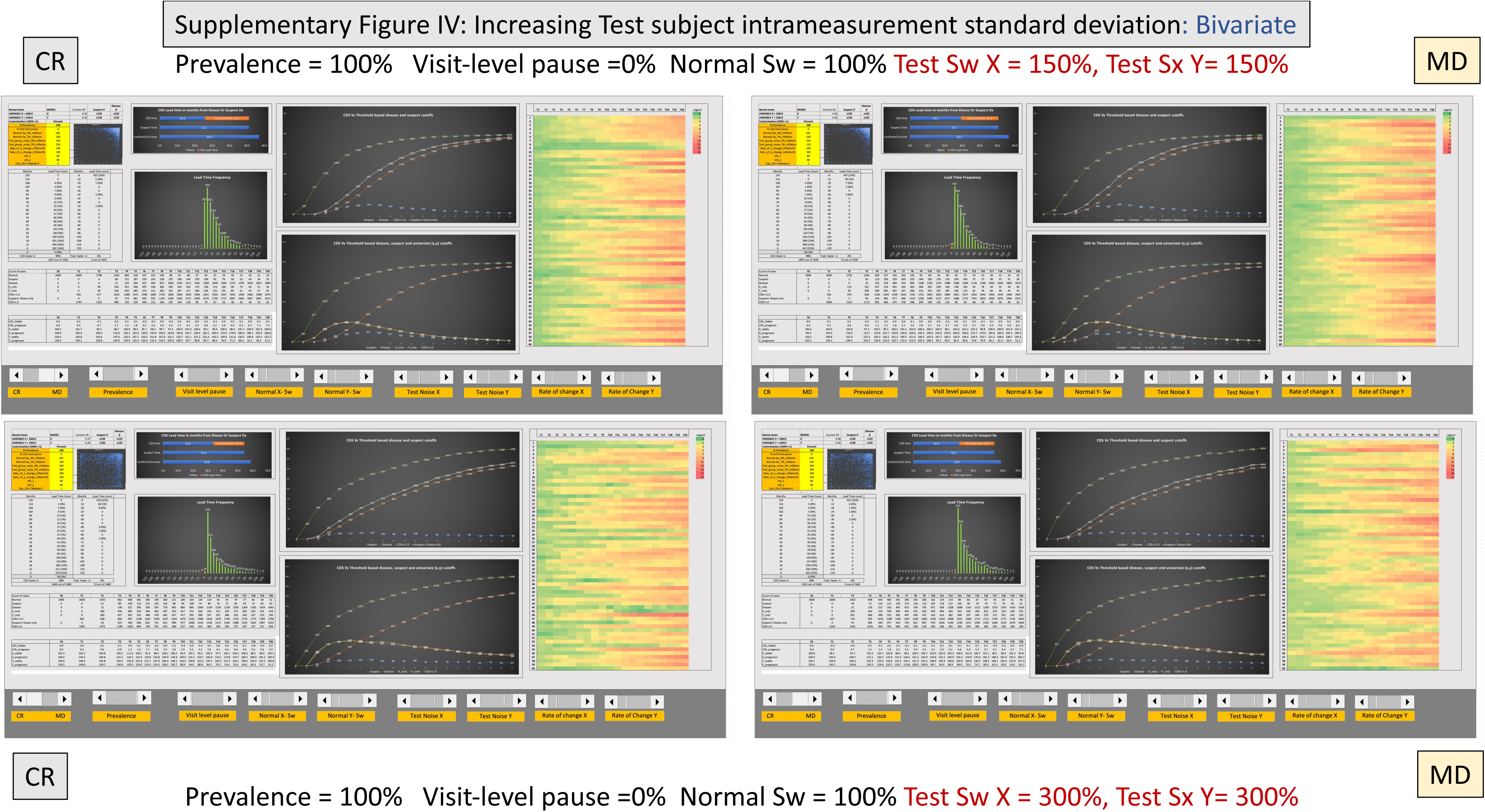
Effect of test eyes intrameasurement standard deviation (Sw) Test group noise was increased to simulate real-world measurement variability. Despite rising Sw to 300%, CDS lead time and drift signal remain intact. Heatmaps show clear clustering, and Mahalanobis-based CDS demonstrates superior resilience. These results suggest the model’s potential to work in noisier-than-usual subject data.

**Supplementary Figure V:**
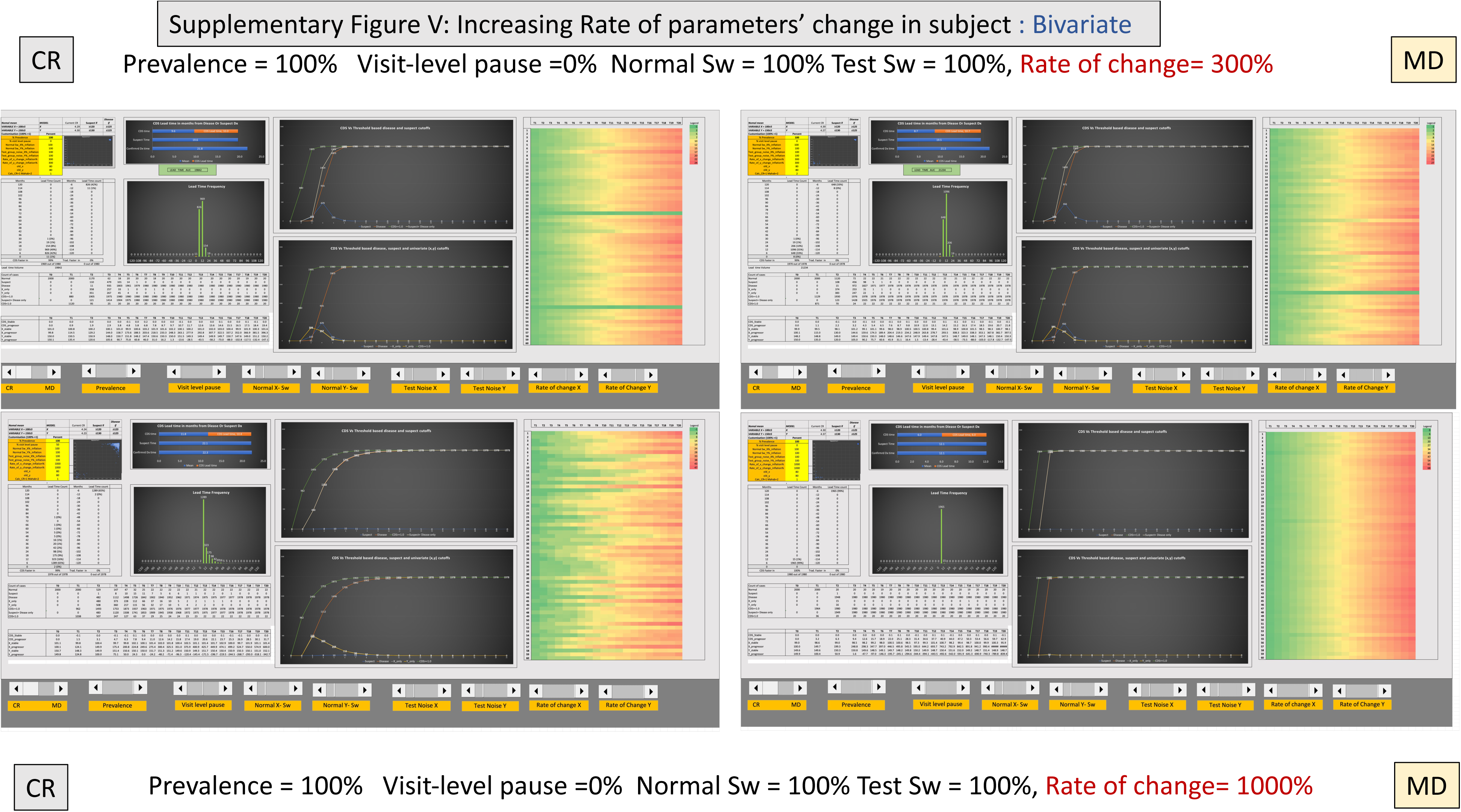
Disease progression speed was increased to test CDS performance under rapid physiological worsening. While all models show reduced lead time advantage at high progression rates (e.g., 1000%), CDS still detects early directional change. This is lines with the biological reality of all classification systems converging in case of increasing disease severity.

