## Appendix A for "Evaluation of a Biplanar Vector-Based Diagnostic Model for Subthreshold Disease: In Silico Stress Testing of Composite Drift Score Performance Under Stochastic, 2D, and 3D Conditions"

**FRAMEWORK DESIGN USED TO ESTABLISH THE PHYSIOLOGICAL AND PATHOLOGICAL PLANE AND THE DERIVED METRICS:**

**Available online at:** Prakash G. Directional drift in biologically meaningful vector planes: a new geometric framework for early disease detection. *medRxiv*. Published June 5, 2025.
doi:10.1101/2025.06.05.25329078

**A. Framework design:**

1. Variable selection and feature space definition: Let $x$ and $y$ denote two biologically meaningful variables reflecting the disease process, which meet the following criteria:
   a) Established clinical relevance in the disease process.
   b) A biologically plausible explanation for their behavior in the disease progression with the presence of directionality
   c) Widely available and commonly used in clinical practice.
   d) Whenever possible, selected variables should be simple, direct measurements or low-complexity derived values with a clear biological rationale.
   e) Low correlation in health subjects

2. Normalization for physiological (normal) subjects: Once identified, the values for these variables are first extracted from a dataset representative of physiological (normal) cases. The two variables are then range normalized:
   a) Physiological Range was calculated as

$$R_{phy} = m_{max} - m_{min} \tag{2(a)}$$

   Where $m \in \{x, y\}$, the original(raw) value of the variable from physiological cases.

   b) If the variable's value is known to increase with disease progression

$$m^{(n)} = \frac{m - m_{min}}{R_{phy}} \quad 2(b)$$

c) If the variable's value is known to decrease with disease progression

$$m^{(n)} = \frac{m_{max} - m}{R_{phy}} \quad 2(c)$$

Thus, normalization is performed according to the transformation rules above resulting in the directionally aligned normalized variables $x^{(n)}$ and $y^{(n)}$

Where: $m \in \{x, y\}$ , and,

- $m \in \{x, y\}$ is the original value of the variable in the physiological cases.
- $m^{(n)}$ is the normalized value for physiological(normal) cases.
- $m_{max}, m_{min}$ are the maximum and minimum values seen in the physiological population.

*Note: Superscripts in parenthesis in this manuscript denote indices (e.g., normalized) and should not be interpreted as exponentiation.*

3. Creation of Physiological plane: This creates a bounded plane defined by normalized coordinates with multiple physiological subjects occupying positions in this coordinate system.

$$P_{phys} = [0,1] \times [0,1] \quad 3(a)$$

$$\left(x^{(n)}, y^{(n)}\right) \in P_{phys} \quad 3(b)$$

Where:

- $P_{phys}$ is the physiological (homeostatic) plane.
- $x^{(n)}, y^{(n)}$ are the normalized values of the two selected variables, each scaled to lie within [0,1].

4. Scaling for pathological (diseased) subjects: The same two variables $x$ and $y$ are extracted from the dataset of patients with early to moderate disease. We exclude patients defined as borderline or suspect to map established disease behavior. Each variable is then scaled by the range noted in the physiological set, using the following transformation rules:

   a) If the variable's value is known to increase with disease progression:

   $$m^{(d)} = \frac{m - m_{min}}{R_{phy}} \tag{4a}$$

   b) If the variable's value is known to decrease with disease progression:

   $$m^{(d)} = \frac{m_{max} - m}{R_{phy}} \tag{4b}$$

   Thus, *scaling (not normalization)* is performed according to the transformation above resulting in the directionally aligned scaled variables $x^{(d)}$ and $y^{(d)}$

   Where: $m \in \{x, y\}$ and,

   - $m \in \{x, y\}$ is the original value of the variable in the pathological cases.
   - $m^{(d)}$ is the *scaled* value.
   - $m_{max}, m_{min}$ are the maximum and minimum values seen in the physiological population as described in the normalization section.
   - $R_{phy}$ is the physiological range, as defined in Equation 2(a)

5. Creation of the pathological plane: in contrast to the physiological plane, this scaled plane extends in the direction of disease progression and is eventually bounded by the End state ($\varepsilon$, the most severe disease state biologically observed before complete loss of structure or function).

$$P_{path} = [0, \varepsilon_x] \times [0, \varepsilon_y] \qquad 5(a)$$

$$\left(x^{(d)}, y^{(d)}\right) \in P_{path} \qquad 5(b)$$

Where:

- $P_{path}$ is the pathological (post-tipping event) plane.
- $\varepsilon_x, \varepsilon_y$ are the scaled disease state maxima derived from the known extreme of the disease, or the End state.
- $x^{(d)}, y^{(d)}$ are the scaled values of the two selected variables.

The scaled disease state maxima ( $\varepsilon_x, \varepsilon_b$) serves only to define the upper edge of the pathological plane; since this study focuses on early drift just beyond the physiological zone, we do not model scenarios where $x^{(d)} \to \varepsilon_x \ or, \ y^{(d)} \to \varepsilon_y$ .

Pathological subjects are conceptualized to have transitioned to this plane once the tipping point is crossed and are now on this plane. They can occupy any location in this feature space and have inter-measurement variability much like the physiological subjects. More importantly, they are expected to move in this coordinate plane towards the direction of worsening pathology unless they have achieved stable disease status showing what we define next as '*meaningful directionality*'.

6. Quantifying '*meaningful directionality*': The first step is identifying ***consistent*** deviation beyond expected oscillation in a subject initially labeled normal. When this deviation is aligned with the direction of disease progression, it can be interpreted as movement characteristic of the pathological plane (or '*meaningful directionality*'). This suggests early disease transition in that subject independent of its distance from the classical, cross-sectional cutoff based, decision boundary. The next step is to quantify this meaningful directionality and create a framework for testing unlabeled cases. For this, we define the following:
    - A pooled noise scalar: $\boldsymbol{\eta}$
    - A pooled disease vector: $\vec{D}$
    - A subject-specific drift vector: $\vec{U}$

a) Noise: The noise is a scalar $\boldsymbol{\eta}$ modeled from the expected physiological noise derived from pooled within-subject standard deviation ($Sw$) over the visits for physiological patients. For this study, one measurement per visit per patient was modeled.

i. For each physiological subject $j \in \{1,2,...,J\}$ the within-subject standard deviation is calculated using all visits $i \in \{1,2,...q_j\}$, where $q_j$ denotes the number of follow-up time points available for subject $j$. The standard deviation for each normalized variable $x^{(n)}, y^{(n)}$ is calculated as:

$$Sw_j{}^{(x^{(n)})} = \sqrt{\frac{\sum_{i=1}^{q_j}\left(x_{j,i}^{(n)} - \bar{x}_j^{(n)}\right)^2}{q_j - 1}} \tag{6(a)}$$

$$Sw_j{}^{(y^{(n)})} = \sqrt{\frac{\sum_{i=1}^{q_j}\left(y_{j,i}^{(n)} - \bar{y}_j^{(n)}\right)^2}{q_j - 1}} \tag{6(b)}$$

$$Cov_j\left(x^{(n)}, y^{(n)}\right) = \frac{1}{q_j - 1}\sum_{i=1}^{q_j}\left(x_{j,i}^{(n)} - \bar{x}_j^{(n)}\right)\left(y_{j,i}^{(n)} - \bar{y}_j^{(n)}\right) \tag{6(c)}$$

Where:

- $x_{j,i}^{(n)}, y_{j,i}^{(n)}$ represent the $i-th$ visit for subject j.
- $\bar{x}_{j,i}^{(n)}, \bar{y}_{j,i}^{(n)}$ are the subject-specific means.

ii. These values were used to compute the joint within-subject standard deviation for every physiological subject *j* as:

$$Sw_j{}^{(x^{(n)},y^{(n)})} = \sqrt{(Sw_j{}^{(x^{(n)})})^2 + (Sw_j{}^{(y^{(n)})})^2 + 2.Cov_j(x^{(n)}, y^{(n)})} \tag{6(d)}$$

1. Final Noise Magnitude: The **coefficient of repeatability** ($CR$), representing the 95% confidence interval for repeated measures for a single variable, is calculated as [63]:

$$CR = 1.96 \cdot \sqrt{2} \cdot Sw \approx 2.77 \cdot Sw \tag{6(e)}$$

In our framework, we extend this concept to the multivariate case, where $Sw$ values are pooled across the two axes. The final scalar noise magnitude is therefore defined as:

$$\boldsymbol{\eta} = 2.77 \cdot \sqrt{\frac{1}{q_j - 1} \sum_{i=1}^{q_j} \left( {Sw_j}^{(x^{(n)}, y^{(n)})} \right)^2} \tag{6(f)}$$

This constructs a radial 95% confidence boundary around expected physiological drift in this joint feature space. While this is conceptually an extension of $CR$ used often to define the agreement between devices, its operational role here differs. It serves as a signal-to-noise denominator in a vector framework thereby providing clinical threshold guidance. This is an extension of its previous application in a univariate case as we used in a previous study [64].

b) Age-related, physiological reference drift: This maps the directionality of the true labeled normal (physiological) cases. Many physiological parameters change with time due to natural aging response and this needs to be mapped to differentiate from pathological change. This can be affected by multiple processes including the age of the subject [65-67]. However, the changes over a shorter period such as less than a decade (compared to entire lifetimes) can be modeled using a linear approximation, especially for exploratory study. This rate of change has been described in multiple longitudinal studies. The resultant vector is:

$$\vec{A} = (x_t^{(a)}, y_t^{(a)}) \tag{6(g)}$$

Where:

$$x_t^{(a)} = \frac{(a_x.t)}{R_{phyx}} \tag{6(h)}$$

$$y_t^{(a)} = \frac{(a_y.t)}{R_{phyy}} \tag{6(i)}$$

With:

$a_x, a_y$ : Age-related drift reference rates for $x, y$

$t$: time between baseline and follow-up

$R_{phyx}, R_{phyy}$: physiological range (raw variables $x$ , $y$) from equation 2(a)

For this exploratory study, we assumed the drift-related change over the 2.5 years we sampled to be negligible. However, the equations were constructed including this drift to keep a placeholder for future iterations of the model.

c) Disease vector: This maps the directionality of the true labeled disease (pathological) cases compared to the physiologically expected (reference) drift. Disease progression can be non-linear over the entire spectrum of pathology. However, the current model is meant for early detection in a population hitherto labeled as normal. The closest approximation for that rate of change seems to be cases with early but established disease (or lower-grade disease) showing progression.

i. For each pathological subject $k \in \{1,2,\dots,K\}$, the scaled change between baseline $(x_{k,0}^{(d)}, y_{k,0}^{(d)})$ and follow-up visit $(x_{k,t}^{(d)}, y_{k,t}^{(d)})$ , for all $t \in T_k$ is calculated adjusting for age-related physiological reference drift ($x_t^{(a)}$, $y_t^{(a)}$). Here$T_k$ denotes the set of valid follow-up time points for subject $k$, and $K$ is the total number of pathological subjects included in the disease vector calculation.

$$\Delta x_{k,t}^{(d)} = \left(x_{k,t}^{(d)} - x_{k,o}^{(d)} - x_t^{(a)}\right) \tag{6(j)}$$

$$\Delta y_{k,t}^{(d)} = \left(y_{k,t}^{(d)} - y_{k,o}^{(d)} - y_t^{(a)}\right) \tag{6(k)}$$

2. Per-subject mean disease trajectory vector: For pathological subject $k$, we compute the average of these vectors across all their follow-up visits:

$$\overline{\Delta x_k^{(d)}} = \frac{1}{T_k}\sum_{t=1}^{T_k} \Delta x_{k,t}^{(d)} \quad 6(l)$$

$$\overline{\Delta y_k^{(d)}} = \frac{1}{T_k}\sum_{t=1}^{T_k} \Delta y_{k,t}^{(d)} \quad 6(m)$$

ii. These subject-level vectors are then averaged to define the **pooled disease vector**.

$$\vec{D} = \left(\overline{\Delta x^{(d)}}, \overline{\Delta y^{(d)}}\right) \quad 6(n)$$

Where:

$$\overline{\Delta y^{(d)}} = \frac{1}{K}\sum_{k=1}^{K} \overline{\Delta x_k^{(d)}} \quad 6(o)$$

$$\overline{\Delta y^{(d)}} = \frac{1}{K}\sum_{k=1}^{K} \overline{\Delta y_k^{(d)}} \quad 6(p)$$

iii. the angle of the pooled disease vector ($\vec{D}$) in reference to the $x, y$ Cartesian frame is:

$$\theta_{\vec{D}} = arctan\left(\frac{\overline{\Delta y^{(d)}}}{\overline{\Delta x^{(d)}}}\right) \quad 6(q)$$

The resulting disease vector ($\vec{D}$) thus serves as the reference direction against which subject-level drift vectors are compared.

d) **Subject drift vector:** For any subject under evaluation (denoted as unknown subject $u$) at follow-up visit $t$ after a baseline visit $t0$ to be evaluated and compared to the pooled metrics of noise and disease vector as above, the following method is performed:

i. Scaling done on raw $variables$ $(x_t, y_t,)$ to create scaled variables for both time $t$ $(x_t^{(u)}, y_t^{(u)})$ and baseline $t0$ $(x_{t0}^{(u)}, y_{t0}^{(u)})$ using transformation principles form Equation 4(a) and 4(b).

ii. The change between scaled values at baseline $(x_{t0}^{(u)}, y_{t0}^{(u)})$ and follow-up $(x_t^{(u)}, y_t^{(u)})$ at time $t$ can be calculated, adjusting for physiological reference drift $(x_t^{(a)}, y_t^{(a)})$ as:

$$\overrightarrow{S_u} = \left(x_t^{(u)} - x_{t0}^{(u)} - x_t^{(a)}, y_t^{(u)} - y_{t0}^{(u)} - y_t^{(a)}\right)$$

6(r)

ii. The magnitude of the subject vector is:

$$\left|\overrightarrow{S_u}\right| = \sqrt{(x_t^{(u)} - x_{t0}^{(u)} - x_t^{(a)})^2 + (y_t^{(u)} - y_{t0}^{(u)} - y_t^{(a)})^2} \quad 6(s)$$

iii. The **angle** $\theta_{\vec{u}}$ of the subject vector is:

$$\theta_{\vec{u}} = \arctan\left(\frac{y_t^{(u)} - y_{t0}^{(u)} - y_t^{(a)}}{x_t^{(u)} - x_{t0}^{(u)} - x_t^{(a)}}\right) \quad 6(t)$$

7. **Metrics to quantify meaningful directionality and establish a decision threshold:**

In the sections above, we constructed the framework of two reference planes: a physiological plane, $P_{phys} = [0,1] \times [0,1])$ , and a pathological plane, $P_{path} = [0, \varepsilon_x] \times [0, \varepsilon_y]$ (see Sections 3a and 5b). We also defined a noise metric $\boldsymbol{\eta}$ from the data in $P_{phys}$ (Eq.6f), a canonical disease vector $\vec{D}$ from data in $P_{path}$ (Eq.6n), a subject-specific drift vector $\overrightarrow{S_u}$ (Eq. 6s) based on scaling principles from $P_{path}$. We now introduce three derived metrics to evaluate a subject's deviation from normal behavior (in magnitude) and alignment toward disease-like behavior:

i. **Change greater than physiological noise**: If a subject has drifted more than that expected by normal physiological oscillation, that becomes the first indicator for concern. Traditionally, limits of repeatability have been applied in a subtractive form (e.g., change > 2 Sw), but such thresholds depend on variable's units (e.g., D (diopters), μm, mg/dl, etc.) and reduce cross variable or cross-domain generalizability. Ratios, on the other hand, are unit agnostic and therefore intuitive to compare. Therefore, we next define the Magnitude to Noise Ratio:

$$\text{MNR}= \frac{|\overrightarrow{S_u}|}{\boldsymbol{\eta}} \qquad 7(a)$$

In its essence, this is a signal-to-noise (SNR) ratio and is an operational metric. Just as a radio-based system uses SNR to deduce a meaningful signal over background static, the MNR intends to deduce subject-level change over the expected noise.

ii. **Directional alignment to disease process:** for the unknown subject $u$ being evaluated, the cosine of the angle between the subject's drift vector $\overrightarrow{S_u}$ and the disease vector $\vec{D}$ is computed as

$$cos(\emptyset_u) = \frac{\vec{D}.\overrightarrow{S_u}}{|\vec{D}|.|\overrightarrow{S_u}|} \qquad 7(b)$$

This value captures the angular alignment between the subject's direction of change and the pooled disease trajectory. For this comparison, a perfectly aligned subject would have $cos(\emptyset_u) = 1$, while a subject drifting orthogonally or in the opposite direction would have values approaching 0 or negative.

Medicine works significantly on pattern recognition and disease phenotypes tend to get replicated in areas of feature space that are proximal to each other. Therefore, we further applied a signed directional weighting. We call this **Directional Emphasis Multiplier**

$$DEM_u = \cos(\emptyset_u).|\cos\emptyset_u| \qquad 7(c)$$

In this study, we used a signed square of the cosine between the subject and disease vector directions. The goal is to produce a curve that favors well-aligned movement and suppresses non-aligned vectors. However, since this metric is customizable, it can be modified based on real-life variations.

iii. The final score combines both directional and magnitude components: The **Composite Drift score** is:

$$CDS_u = DEM_u \cdot MNR_u \quad 7(d)$$

This scalar, unitless value reflects both the strength and directionality of pathological drift and forms the basis for early detection in this framework. It is intuitively set at a threshold of ≥1.0 (perfect alignment with disease process and change greater than seen with noise, which can vary over multiple case scenarios as we discuss later in results and discussion).

8. **Expansion to multidimensional space and adjustments for high covariance**: For a ≥3-dimensional model, or in cases where there is a significant covariance, the above method may be challenging even after scaling. We explored the candidate methods for such situations. We evaluated Mahalanobis distance (MD) as a prospective alternative method to set up this for future iterations of this model [68-70]. CR is used more commonly in repeatability studies and therefore it is more intuitive for clinicians. Mahalanobis distance is used as a robust compensation for the covariance in multiple fields including medicine The two parameters, we used in this study to represent $x$ *and* $y$ , Kmax and TCT do not have significant covariance in normal cases. However, we suspect that this can be a potential concern in some clinical situations, especially when this framework is expanded. Ideally, matrix manipulation is easier in a programming environment such as Python, however, in this exploratory study we wanted to ensure the generalizability and have used Excel environment. In next steps, we outline the method we used to calculate Mahalanobis distance:

i. For each normal subject j ∈ $\{1,2,...,J\}$, and for each follow-up point $t \in T_j$ we used the normalized values obtained from Equations 2a,2b and computed the change relative to baseline for variables $x$ and $y$ respectively. These were arranged in row vectors:

$$M_{j,i} = \left[\partial x_{j,t}^{(n)} \quad \partial y_{j,t}^{(n)}\right] \qquad 8(a)$$

Where:

$\partial x_{j,t}^{(n)}$ is the change in normalized $x$ compared to baseline for subject $j$ at follow-up visit $t$

$\partial \mathrm{y}_{j,t}^{(n)}$ is the change in normalized $y$ compared to the baseline for subject $j$ at the follow-up visit $t$

*(It should be emphasized that this method does not use the baseline visit as a separate visit like when calculating Sw but the difference of baseline from follow-ups).*

ii. Aggregating across all subjects and all follow-up time points yielded the following data matrix with all the data points.

$$M \in \mathbb{R}^{Nx2} \qquad 8(b)$$

Where:

N=$\sum_{j=1}^{J} T_j$

iii. From this matrix $M \in \mathbb{R}^{Nx2}$ we calculated the sample covariance matrix:

$$\Sigma = Cov(V) \in \mathbb{R}^{2x2} \quad = \quad \begin{bmatrix} Var_x & Covar_{x,y} \\ Covar_{x,y} & Var_y \end{bmatrix} \qquad 8(c)$$

This matrix contains variance of $x$ and $y$ along the diagonal and covariance between $x$ and $y$ off-diagonal.

iv. For each subject under evaluation (denoted as unknown subject $u$ ) at follow-up visit $t$, we define their normalized drift vector relative to baseline as:

$$v_{u,t} = \begin{bmatrix} \partial x_{u,t}^{(s)} \\ \partial y_{u,t}^{(s)} \end{bmatrix} \qquad 8(d)$$

Where:

$\partial x_{u,t}^{(s)}$ is the change in scaled $x$ compared to baseline for subject $u$ at follow-up visit $t$

$\partial y_{u,t}^{(s)}$ is the change in scaled $x$ compared to baseline for subject $u$ at follow-up visit $t$

(scaling done as in equation 4a,4b, as the status (disease or normal) is unknown)

v. The Mahalanobis distance (MD) for the change seen in subject $u$ at follow-up $t$ from the pool of expected change in normal subjects is therefore calculated as:

$$MD_{u,t} = \sqrt{v_{u,t}^{T} \Sigma^{-1} v_{u,t}} \qquad 8(e)$$

Where:

$v_{u,t}^{T}$ is the transpose of the matrix $v_{u,t}$ for subject vector from equation 8(d)

$\Sigma^{-1}$ is the inverse of the covariance matrix ($\Sigma$)of pooled normal subjects from equation 8(c)

The next step is to compare this distance to a reference point. As a guideline, most studies including ours have used 2.77 times $Sw$ as $CR$ ( see equation 6e), which translates into 95% of the observed change within expected variation in a univariate model. However, Mahalanobis distance follows a chi-square distribution. Therefore, we utilize the chi-square distribution for 2 degrees of freedom to derive an equivalent multivariate confidence boundary. The 95th and 99$^{th}$ percentile of this distribution corresponds to approximately 5.99 and 9.21. The square root of these values gives thresholds as below:

$$\sqrt{5.99} \approx 2.45 \quad 8(f)$$

$$\sqrt{9.21} \approx 3.03 \quad 8(g)$$

vi. To compare with our Euclidean distance-based CR model (from equation 6f) we used the scaling logic derived above. We used 2.45 for the initial iteration as this corresponds approximately to the 95% percentile logic used for the CR. This will change based on the degrees of freedom and required alpha and is customizable for future iterations. To keep intuitive comparability with the MNR derived above in 7a, we call this entity.

$$MNR_u^{(MD)} = \frac{MD_{u,t}}{2.45} \quad 8(h)$$

vii. The CDS hence calculated based on this $MNR_u^{(MD)}$ is:

$$CDS_u^{(MD)} = DEM_u \cdot MNR_u^{(MD)} \quad 8(i)$$

Where:

$DEM_u$ is the same Directional Emphasis Multiplier as in 7(c)
